# Heterogenous Cellular and Humoral Immune Trajectories after SARS-CoV-2 Infection: Compensatory Responses in a Population-Based Cohort

**DOI:** 10.1101/2021.12.15.21267776

**Authors:** Dominik Menges, Kyra D. Zens, Tala Ballouz, Nicole Caduff, Daniel Llanas-Cornejo, Hélène E. Aschmann, Anja Domenghino, Céline Pellaton, Matthieu Perreau, Craig Fenwick, Giuseppe Pantaleo, Christian R. Kahlert, Christian Münz, Milo A. Puhan, Jan S. Fehr

## Abstract

To better understand the development of immunity against SARS-CoV-2 over time, we evaluated humoral and cellular responses a population-based cohort of SARS-CoV-2-infected individuals covering the full spectrum of COVID-19 up to 217 days after diagnosis. We characterized anti-Spike (S)-IgA and -IgG antibody responses in 431 individuals and found that about 85% develop and maintain anti-S-IgG responses over time. In a subsample of 64 participants selected for a detailed characterization of immune responses, we additionally evaluated anti-Nucleocapsid (N)-IgG antibodies and T cell responses specific to viral Membrane (M), N, and S proteins. Most participants had detectable T cell responses to at least one of the four peptide pools analyzed, which were more frequent than antibody seropositivity. We found a moderate correlation between antibody and T cell responses, which declined over time and suggests important variability in response patterns between individuals. The heterogeneity of immune trajectories was further analyzed using cluster analyses taking into account joint antibody and T cell responses over time. We identified five distinct immune trajectory patterns, which were characterized by specific antibody, T cell and T cell subset patterns along with disease severity and demographic factors. Higher age, male sex, higher disease severity and being a non-smoker was significantly associated with stronger immune responses. Overall, the results highlight that there is a consistent and maintained antibody response among most SARS-CoV-2-infected individuals, while T cell responses appear to be more heterogenous but potentially compensatory among those with low antibody responses.

**One Sentence Summary:** Presence of heterogenous immune response trajectories after SARS-CoV-2 infection with potential compensatory role of T cells among individuals with low antibody responses.

## INTRODUCTION

Almost two years after its start, the SARS-CoV-2 pandemic remains a threat to public health worldwide and has resulted in hundreds of million cases and millions of deaths globally (*1*). Control of the pandemic now relies largely on the development of robust immunity in the population after infection, vaccination, or both, which necessitates an in-depth understanding of humoral and cellular responses to the virus.

Several studies have characterized B and T cell-mediated immune responses against SARS-CoV-2 and showed that both antibodies and T cells are generated in most people after infection (*2–19*). Immunoglobulin (Ig) spike specific antibodies have been detected within a few days after infection (*4, 14–16, 19–22*) and their neutralizing capacity has been confirmed (*5, 12, 14, 18, 22–26*), with the magnitude of response positively correlating with disease severity (*7, 17, 21, 27–30*). However, studies have also shown varying results regarding the longitudinal changes of these antibody responses. While certain studies found that antibodies persist for several months after infection (*8, 10, 25, 26, 29, 31–37*), some reported declining levels of antibodies within a few months, particularly among those with mild disease (*30, 38–42*), raising concerns about the longevity of protection against re-infection. On the other hand, T cell-mediated immunity generally seems to be more stable. There is some evidence that robust T cell responses are developed even after mildly symptomatic coronavirus disease 2019 (COVID-19) or asymptomatic infection and despite low levels or a complete absence of antibodies (*8–10, 28, 31, 43, 44*). While these studies have significantly advanced the general understanding of immunological responses after SARS-CoV-2 infection, most were conducted in highly specific populations or using convenience samples. Few have longitudinally assessed the diverse components of the immune response within the same individuals (*2, 8, 28, 31*) and in cohorts representative of the entire range of the infected population (*8, 28, 31*). Furthermore, despite evidence of heterogeneous immune responses between individuals and the knowledge that antibodies and T cells act together at different stages of a viral infection to protect against severe disease and re-infection, little attention has been paid to capturing or describing the diverse joint trajectories of antibodies and T cells within individuals infected with SARS-CoV-2.

In this study, we analyzed longitudinal patterns of humoral and cellular immune responses for up to six months post-diagnosis in a population-based cohort of SARS-CoV-2-infected individuals across the full COVID-19 disease spectrum. We first characterized anti-Spike (S) IgA and IgG antibody responses and estimated their decay rates up to 217 days post-diagnosis in 431 individuals. In a subsample of 64 participants selected to cover the full clinical spectrum of SARS-CoV-2 infection and antibody responses, we performed a detailed evaluation of anti-Nucleocapsid (N) IgG antibodies and T cell responses specific to viral Membrane (M), N, or S proteins (including the S1 and the majority of the S2 domains). Based on the co-evolution of both antibody and T cell responses over time, we defined five distinct immune trajectory patterns post-infection. Overall, this longitudinal study highlights that there is a consistent antibody response among most, but not all, SARS-CoV-2-infected individuals up to six months, while T cell responses appear to be more heterogenous, but potentially compensatory among those with low antibody responses.

## RESULTS

### Study population and sample measurements

In this analysis, we included measurements from 431 SARS-CoV-2-infected individuals, selected as a random, age-stratified sample of all SARS-CoV-2-infected individuals reported to the Department of Health of the Canton of Zurich between 06 August 2020 and 26 January 2021. Peripheral blood samples were collected for immunological analysis at two weeks, one month, three months, and six months after diagnosis of infection (Fig. 1A). The median age of participants was 52 years (interquartile range (IQR) 35–68 years) and 212 (49%) were female (Supplementary Table S1). Fifty-nine (14%) of the participants were smokers and 131 (30%) had at least one comorbidity, most commonly hypertension (16%) and respiratory diseases (7%). Most participants (83%) were symptomatic, with 163 (38%) reporting between one and five symptoms and 192 (45%) reporting six symptoms or more during acute infection. Within two weeks of diagnosis, 18 (4%) participants required hospitalization for reasons related to COVID-19, among which two participants were admitted to an intensive care unit. At six months after diagnosis, three participants (0.7%) reported a SARS-CoV-2 reinfection and 80 (19%) were vaccinated with at least one dose (all with mRNA vaccines).

**Fig. 1.**
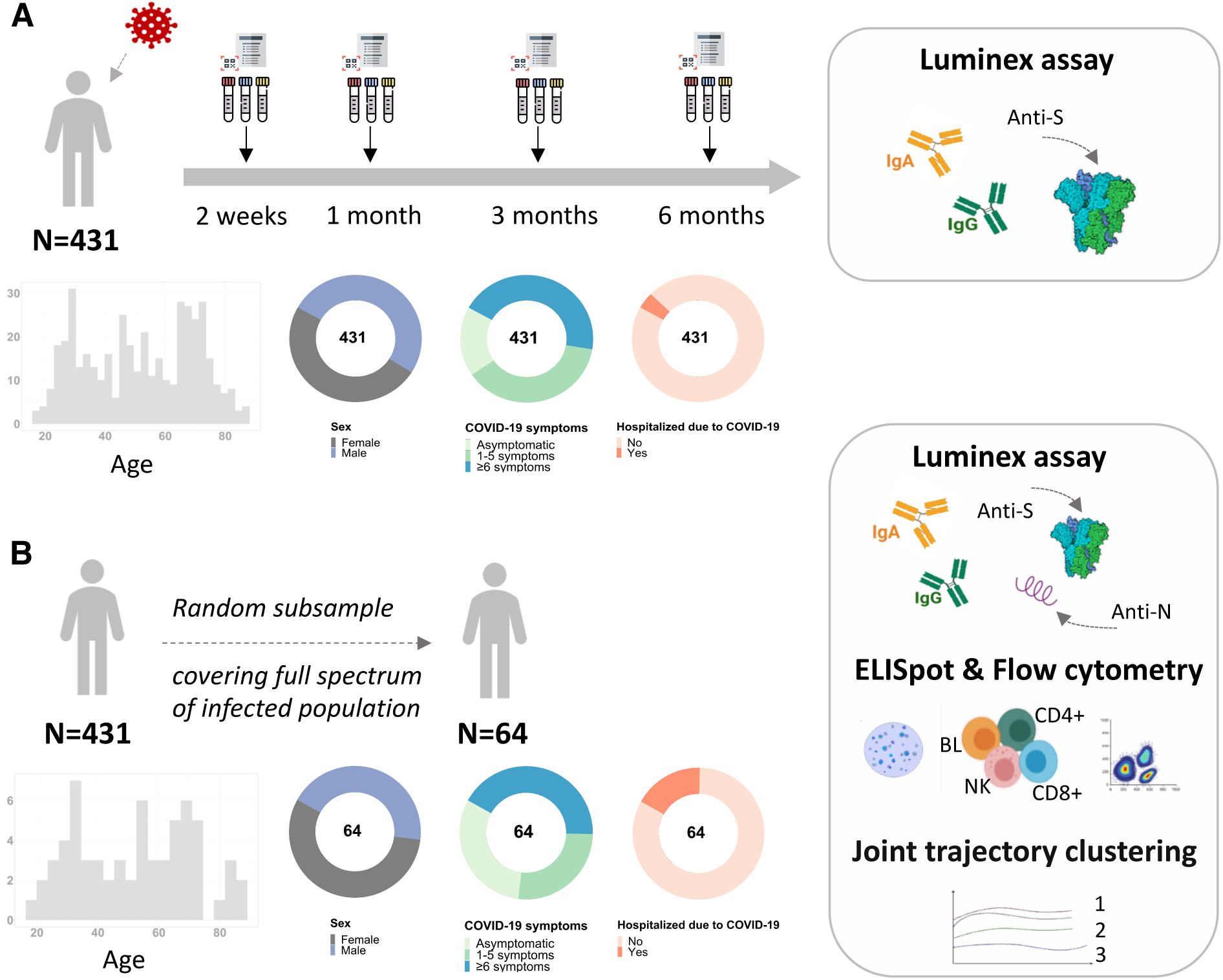
Study design and population characteristics. (**A**) Figure showing the overall study design, assessments conducted on and baseline clinical characteristics of all enrolled participants (n=431). (**B**) Figure showing the assessments conducted on and clinical characteristics of a randomly selected subsample of participants covering the full range of the infected population (n=64).

We selected a subsample of 64 individuals for a detailed characterization of immune responses (i.e., additional anti-N antibody testing as well as ELISpot and flow cytometric analyses of virus-specific T cells), aiming to cover the full spectrum of disease severity (i.e., asymptomatic to hospitalized), antibody responses (i.e., low to high anti-S-IgA and -IgG antibody responses) and balanced in terms of age and sex (Fig. 1B). In this subsample, the median age was 53.5 (IQR 33.5–68 years) and 56% of participants were female (Supplementary Table S1). 69% of the participants had symptoms of COVID-19, with 27% reporting between one and five symptoms and 42% reporting six or more symptoms. Eleven (17%) of the participants were hospitalized due to COVID-19 and one participant required admission to an intensive care unit.

In summary, anti-S antibody responses were characterized in 431 SARS-CoV-2 infected individuals at two weeks, one month, three months and six months after diagnosis. Anti-N antibody responses, as well as M-, N-, S1 domain- and S2 domain-specific T cell responses were additionally measured in a subsample of 64 individuals, covering the full clinical spectrum and range of S-specific antibody responses at these timepoints.

### Longitudinal follow-up of patients recovering from COVID-19 highlights sustained anti-Spike IgG responses, anti-Nucleocapsid and anti-Spike Domain 2 T cell responses, and CD8^+^ T cell responses

Using a highly sensitive Luminex-based assay (*45*), we first assessed levels of SARS-CoV-2 S-specific IgA and IgG or N-specific IgG in blood plasma over time. Within the full study population (n=431) we found that the median anti-S-IgA response was highest by two weeks post-diagnosis, after which responses declined steadily up to six months (Fig. 2A, Supplementary Tables S2 and S3). In contrast, anti-S-IgG responses peaked later at one month post-diagnosis, persisted until three months, and then waned slightly until the six month follow-up (Fig. 2B). Among the subsample (n=64) we found that anti-N-IgG responses followed a similar initial kinetic to that of anti-S-IgG (peaking at month one), but the decline over time was more pronounced (Fig. 2C). From these data, we utilized a linear decay model adjusted for time from diagnosis to maximum antibody concentration, to estimate the half-life of each antibody subtype. From the full population we estimated the half-life of anti-S-IgA to be 71 days (95% CI 66–76 days, Fig. 2D) and 145 days (135–156 days; Fig. 2E) for anti-S-IgG. From the subsample, we estimated the half-life of anti-N-IgG to be 86 days (76–99 days; Fig. 2F).

**Fig. 2.**
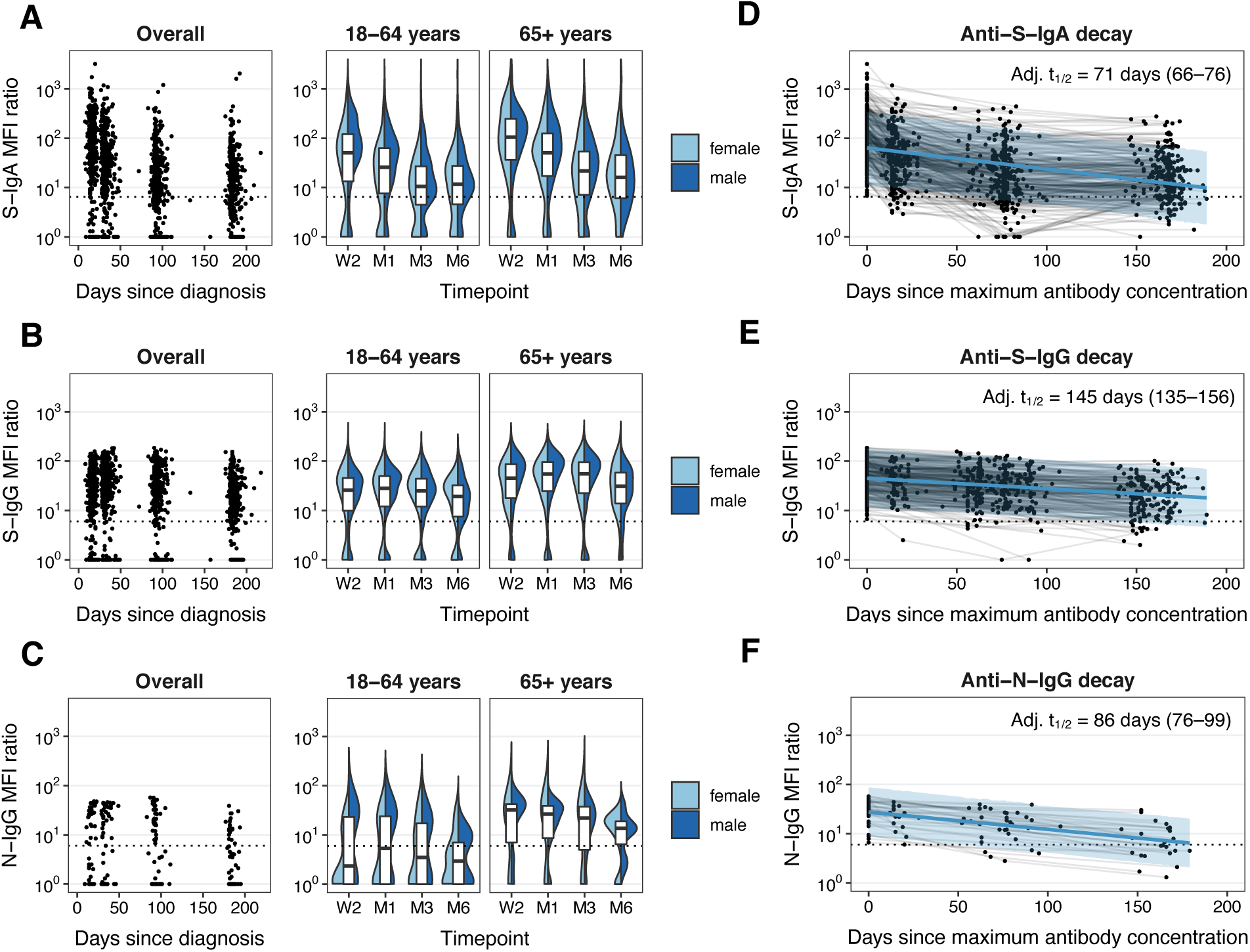
Anti-SARS-CoV-2 S-IgA, S-IgG, N-IgG-specific antibody responses over time. (**A**) Scatter and distributional plots demonstrating the measured mean fluorescence intensity (MFI) ratios for anti-S-IgA antibodies in the overall study population (n=431), overall and stratified by age groups and sex. MFI: mean fluorescence intensity, W2: two weeks, M1: one month, M3: three months, M6: six months after diagnosis. (**B**) Scatter and distributional plots of measured MFI ratios for anti-S-IgG antibodies in the overall study population (n=431), overall and stratified by age groups and sex. (**C**) Scatter and distributional plots of measured MFI ratios for anti-S-IgG antibodies within the subsample of individuals undergoing detailed testing (n=64), overall and stratified by age groups and sex. (**D**) Anti-S-IgA antibody decay estimation based on mixed linear regression model. Adj. t_1/2_: half-life based on model adjusted for time from diagnosis to maximum MFI ratio, age group, sex and symptom count, using a random intercept for each individual in the study. (**E**) Anti-S-IgG antibody decay estimation based on mixed linear regression mode. (**F**) Anti-N-IgG antibody decay estimation within the subsample, based on mixed linear regression model.

We further evaluated the level of seropositivity for each antibody subtype. In the full study population, we found that 83% (79–86%) and 82% (78–86%) of individuals were anti-S-IgA or anti-S-IgG seropositive, respectively at two weeks post-diagnosis (Supplementary Fig. S1A, Supplementary Table S2). While the percentage of anti-S-IgA-seropositive participants decreased over time reaching 70% (65–75%) at six months, the percentage remaining anti-S-IgG-seropositive was more stable (81%, 77–85%). Overall, 85% of the population was positive for either anti-S-IgA or -IgG at three (81–88%) and six (81–88%) months post-diagnosis. Of note, only ten participants (2.3%) were initially seropositive (either anti-S-IgA or -IgG) but became seronegative within six months. Estimates were similar in age group-weighted sensitivity analyses (Supplementary Table S4). In the subsample, 59% of the participants were seropositive each for anti-S-IgA and -IgG, and 54% were seropositive for anti-N-IgG at two weeks. At six months, 61%, 66% and 41% were seropositive for anti-S-IgA, anti-S-IgG and anti-N-IgG, respectively (Supplementary Fig. S1B, Supplementary Table S3). Taken together, these findings suggest that S-specific IgG responses are more robust than S-specific IgA or N-specific IgG responses following SARS-CoV-2 infection.

In the subsample, we additionally evaluated virus-specific T cell populations using an interferon-gamma ELISpot assay, assessing responses to overlapping peptide pools spanning the entire SARS-CoV-2 M and N proteins, the S1 domain of the S protein (which contains the Receptor Binding Domain (RBD)), or a mix of other predicted immunodominant epitopes within the S protein, containing the majority of the S2 domain (which we refer to here as S2 for simplicity). We found that 84% (72–91%) of individuals had detectable T cell responses to at least one of the four tested peptide pools at two weeks post-infection, dropping to 71% (58–81%) at six months (Fig. 3A, Supplementary Fig. S1C). Positivity to individual peptide pools ranged from 71% (59–82%; S2) to 80% (68–89%; M) at two weeks to 46% (33–59%; S1) to 51% (38–64%; N) at six months, with no discernible differences in positivity between pools at any timepoint (Fig. 3A, Supplementary Fig. S1C, Supplementary Table S3). Assessing the combined T cell response to all four peptide pools over time, M-specific T cells made up 33% of the total detectable T cell response at two weeks of follow-up, compared to 29% S1-specific, 21% N-specific, and 16% S2-specific T cells (Fig. 3B). Proportions remained stable up to six months, contributing 26%, 26%, 27% and 21% to the total T cell response, respectively. Similar to antibody responses, we estimated decay kinetics for individual peptide pool-specific T cell responses or for the overall T cell response by summing the responses to individual peptide pools. For the overall T cell response, we estimated a half-life of 145 (80–781) days (Supplementary Fig. S1D). For individual peptide pools, we estimated the half-life of M and S1-specific T cells to be substantially shorter (128 days and 126 days, respectively) compared to that of N and S2-specific T cells (227 days and 266 days, respectively), suggesting that the latter may provide more durable responses following infection (Supplementary Fig. S1E to S1H).

**Fig. 3.**
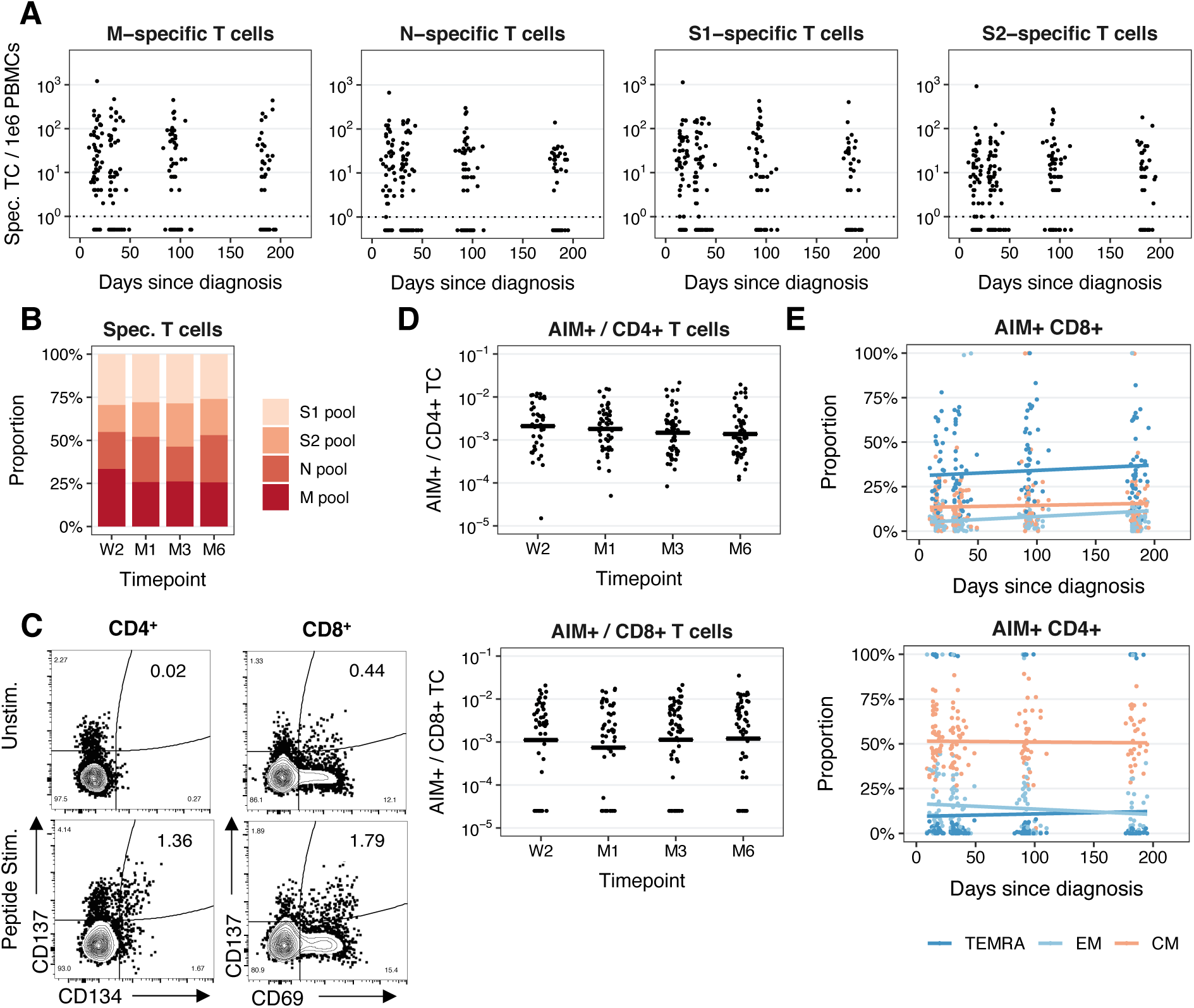
M-, N-, S1- and S2-specific T cell responses over time. (**A**) Number of T cells responding to M, N, S1 or S2 overlapping peptide pools per 1e6 PBMCs by day post-diagnosis. (**B**) Fraction of the total T cell response specific for each peptide pool at indicated timepoints post-diagnosis. Spec. TC: specific T cells, W2: two weeks, M1: one month, M3: three months, M6: six months after diagnosis. (**C**) Representative flow cytometry plots depicting AIM^+^ (CD134^+^CD137^+^) CD4^+^ (left) and (CD69^+^CD137^+^) CD8^+^ (right) populations in unstimulated (top) or pooled SARS-CoV-2 peptide-stimulated (bottom) PBMCs. (**D**) AIM^+^ cells as a fraction of CD4^+^ or CD8^+^ T cells at indicated timepoints post-diagnosis. Horizontal lines represent mean at timepoint. (**E**) Percentage of CD4^+^ or CD8^+^ AIM^+^ T cells with TCM (CD45RA^−^CCR7^+^), TEM (CD45RA^−^CCR7^−^) or TEMRA (CD45RA^+^CCR7^−^) phenotypes by day post-diagnosis.

We further evaluated SARS-CoV-2-specific CD4^+^ and CD8^+^ T cells in the subsample using a Flow Cytometry-based Activation-Induced Marker (AIM) assay. Using the markers CD137 (4-1BB) and CD134 (OX-40) for CD4^+^ T cells or CD69 and CD137 for CD8^+^ T cells, we determined frequencies of activated cells following in vitro stimulation with a combined pool of M, N, S1, and S2 peptides (Fig. 3C). We found similar frequencies of total peripheral blood AIM^+^CD4^+^ or AIM^+^CD8^+^ T cells (approximately 0.2–0.3%) at two weeks post-diagnosis. For CD4^+^ T cells, this declined over time to 0.1% at six months, although this did not reach statistical significance (p=0.11, Kruskal-Wallis test; Fig. 3D). In contrast, for CD8^+^ T cells, frequencies of AIM^+^ cells were more stable over time, remaining at 0.2% up to six months post-infection. In assessing the phenotype of AIM^+^ T cells in the blood, CD4^+^AIM^+^ T cells were predominantly central memory T cells (TCM), while CD8^+^AIM^+^ T cells were predominantly T effector memory cells re-expressing CD45RA (TEMRA; Fig. 3E). Taken together, our results suggest that, CD8^+^ T cells, in addition to anti-S IgG, and M- and S-specific T cells, act as more long-lasting components of SARS-CoV-2-specific immunity following infection.

### Sustained, strong concordance within antibody or T cell responses, but moderate concordance between antibody and T cell responses which declines over time

To better understand the relationship within and between antibody and T cell responses over time, we next used the subsample to evaluate concordance by comparing the levels of antibody and T cell responses. Within the overall antibody response (anti-S-IgA, -S-IgG, -N-IgG), we observed a strong, positive correlation in the magnitude of response among different subtypes at each timepoint post-diagnosis (Fig. 4A). Similarly, within the overall T cell response, we observed a strong, positive correlation in the magnitude of response among different peptide pools at each timepoint (Fig. 4A). In comparing between antibody and T cell responses, however, the correlation was less robust, but still present. At two weeks post-diagnosis, we observed a weak to moderate, positive correlation between the magnitude of each antibody subtype evaluated and overall (pooled M, N, S1, S2) T cell responses (anti-S-IgA Spearman r=0.38; anti-S-IgG Spearman r=0.37; anti-N-IgG Spearman r=0.47; Fig. 4A). This correlation decreased over time and was weak or no longer observed by six months of follow-up (anti-S-IgA Spearman r=0.16; anti-S-IgG Spearman r=0.25; anti-N-IgG Spearman r=0.24). These findings highlight that antibody and T cell responses tend to behave similarly (in terms of magnitude) early in the immune response, but not necessarily in the longer term. Furthermore, the magnitude of the response by one antibody subtype is better predicted by the response to other antibody subtypes than by T cell responses, and vice versa.

**Fig. 4.**
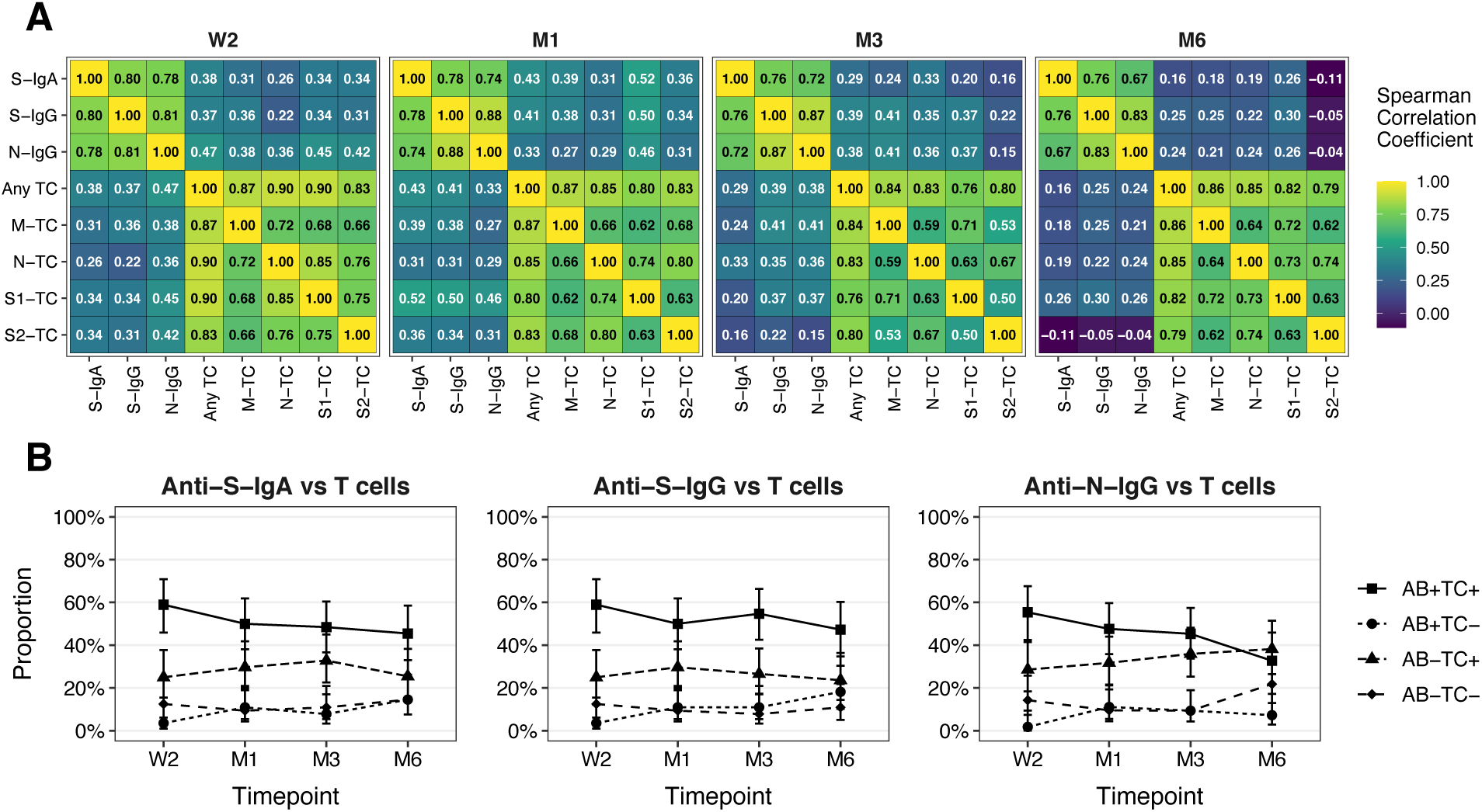
Relationship between antibody and T cell responses over time. (**A**) Heatmaps demonstrating the correlation between anti-S-IgA and IgG and anti-N-IgG antibody subtypes and M-, N-, S1- and S2-specific T cells at the different timepoints. Numbers in individual cells correspond to the Spearman correlation coefficient. W2: two weeks, M1: one month, M3: three months, M6: six months after diagnosis. (**B**) Panel demonstrating the proportion of participants with concordant and discordant results between anti-S-IgA, anti-S-IgG and anti-N-IgG antibody subtypes and overall T cell positivity over time.

We also assessed the proportion of individuals that were “positive” or “negative” for antibody and T cell responses at each timepoint, comparing anti-S-IgA, anti-S-IgG, or anti-N-IgG to pooled T cells responses (positive to any peptide pool; Fig. 4B), or to specific peptide pools (Supplementary Fig. S2A). At two weeks post-diagnosis, approximately 55–60% of individuals were both antibody “positive” and pooled-T cell “positive”, while approximately 10-15% were both antibody “negative” and pooled-T cell “negative” (Fig. 4B). Thus, overall percent concordance between antibody responses and pooled T cell responses was approximately 70%, which was similar for anti-S-IgA, anti-S-IgG, and anti-N-IgG. By six months, concordance between pooled T cell responses dropped to 55–60% (depending on antibody subtype). For both anti-S-IgA and anti-S-IgG, this appeared to be primarily due to an increased fraction of participants becoming T cell “negative”. For anti-N-IgG, this drop appeared to be additionally driven by an increasing fraction of anti-N-IgG “negative” participants. Patterns of antibody subtype and T cell concordance were also similar between T cells specific to individual peptide pools (M, N, S1, S2), and analogous trends were observed when evaluating test agreement using Cohen’s Kappa (Supplementary Fig. S2A and S2B). Together, these findings suggest that SARS-CoV-2-specific antibody and T cell responses correlate early after infection, but that this correlation decreases with increasing time after infection.

### Clustering of antibody and T cell responses reveals five distinct joint immune trajectories

We subsequently explored whether distinct patterns of joint antibody and T cell response trajectories could be observed using a non-parametric longitudinal clustering algorithm (*46, 47*). We identified five distinct joint trajectories of antibody subtypes and peptide pool-specific T cell responses within the subsample (Fig. 5A and 5B). Clusters of participants were primarily defined by the presence (clusters 1-4) or absence (cluster 5) of antibody responses, as well as distinct T cell trajectories. When present, antibody trajectories generally followed the decay patterns observed in the overall study population, characterized by waning anti-S-IgA and anti-N-IgG as well as persistent anti-S-IgG. Meanwhile, T cell trajectories between clusters were more heterogenous. We additionally examined T cell subsets using data from flow cytometric analyses, which did not influence clustering, to identify differences in immune phenotypes between clusters.

**Fig. 5.**
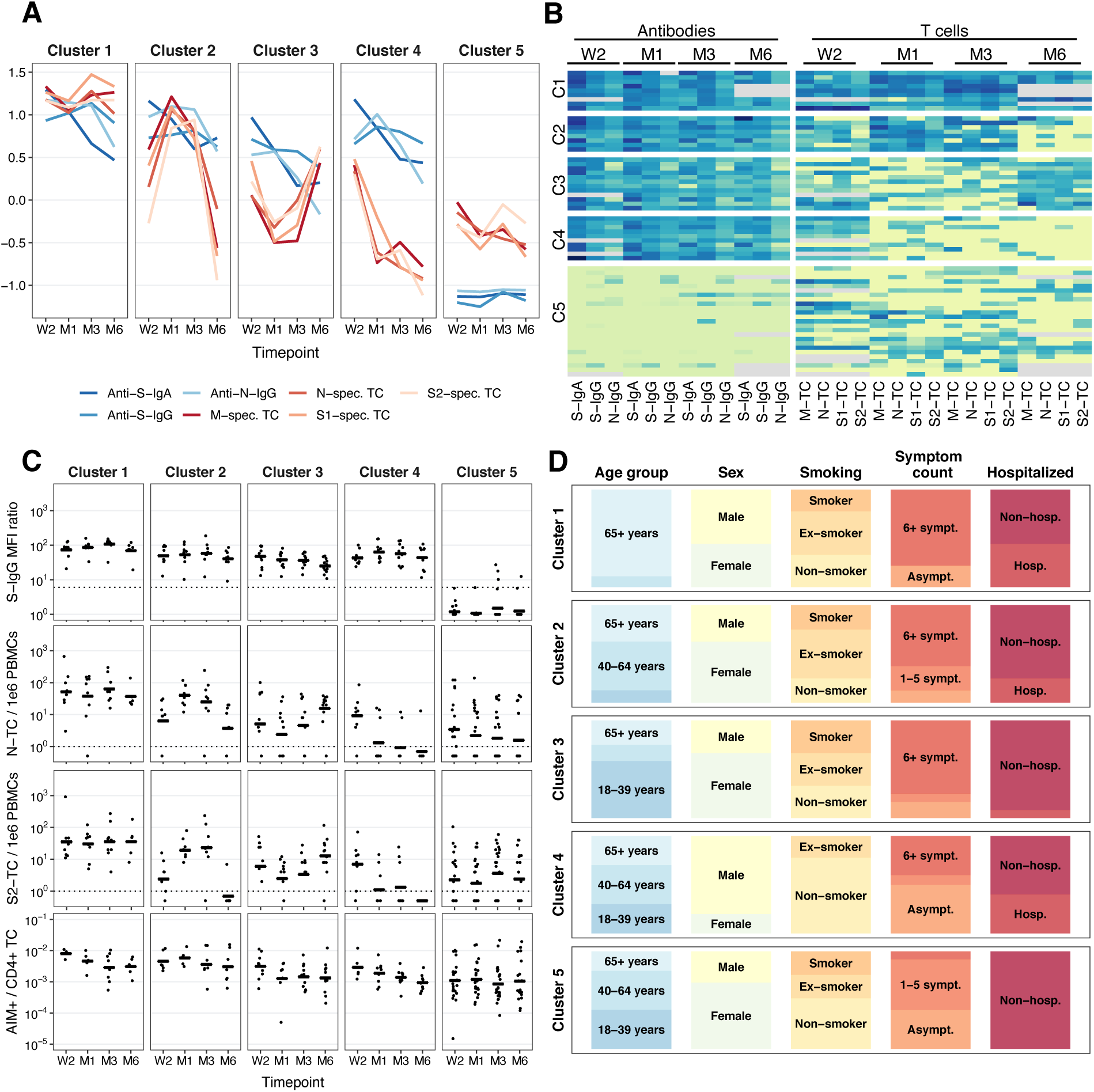
Clustering of antibody and T cell response trajectories. (**A**) Panel demonstrating the mean MFI ratios of anti-S-IgA, anti-S-IgG and anti-N-IgG antibodies and mean M-, N-, S1-, and S-specific T cells/1e6 PBMCs in each of the five clusters over time. Displayed data was natural logarithm-transformed and normalized. W2: two weeks, M1: one month, M3: three months, M6: six months after diagnosis. (**B**) Heatmap showing the MFI ratios of anti-S-IgA, anti-S-IgG and anti-N antibodies and M-, N-, S1-, and S-specific T cells/1e6 PBMCs for all participants belonging to the identified clusters 1 to 5. Gray color indicates missing values. (**C**) Dot plots of anti-S-IgG MFI ratio, N-specific T cells/1e6 PBMCs, S-specific T cells/1e6 PBMCs and frequency of AIM^+^ of CD4^+^ T cells in the five clusters over time. Horizontal lines represent mean at timepoint. (**D**) Bar plot showing the proportion of participants according belonging to different age groups (18-39 years, 40-64 years, ≥65 years), sex (male and female), smoking status (non-smoker, ex-smoker, smoker), number of symptoms reported (asymptomatic, 1-5 symptoms, ≥6 symptoms), and hospitalization status (non-hospitalized, hospitalized) during acute infection. Asymp.: asymptomatic, symp.: symptoms, hosp.: hospitalized.

The *first cluster* (14% of subsample participants) was characterized by high antibody and T cell responses, which remained high across all evaluated timepoints. T cell response trajectories were similar for M, N, S1, S2 peptide pools, peaking at three months with a subsequent slight decline up to six months post-diagnosis. N- and S2-specific T cell responses were the most robust; at six months post-diagnosis, 88% participants had detectable N-specific T cells and 75% had S2-specific T cells (Fig. 5C, Supplementary Fig. S3A). By flow cytomatric analysis, individuals in cluster 1 had the highest total CD4^+^ T cell count at one and three months post-diagnosis, but showing a decline at six months (Supplementary Fig. S3B). Additionally, total CD8^+^ T cell numbers were lower compared to clusters 3-5 and showed a decline from three to six months. We further found that individuals in this cluster had higher frequencies of both CD4^+^AIM^+^ and CD8^+^AIM^+^ virus-specific T cells at all timepoints compared to all other clusters except cluster 2 (Fig. 5C). We noted that participants belonging to the first cluster were mostly older than 65 years (89%), male (56%) and 22% were smokers (Fig. 5D). The majority (78%) had more than six COVID-19 symptoms and 44% were hospitalized within two weeks of diagnosis, indicating more severe disease. Thus, cluster 1 tended to represent older individuals with more severe disease and also robust antibody and cell-mediated immune responses.

The *second cluster* (12% of the subsample) was characterized by persistently high anti-S-IgA and -IgG responses in all participants, who also demonstrated a steep increase in virus-specific T cells from two weeks to one month post-diagnosis. T cell positivity subsequently declined until six months post-diagnosis, with N- and S2-specific T cells being present in 63% and 13% of participants, respectively (Fig. 5C, Supplementary Fig. S3A). By flow cytometry, we found that individuals in this cluster had lower total CD8^+^ T cell numbers than those in all other clusters at two weeks and one months, increasing up to three months post-diagnosis (Supplementary Fig. S3B). Meanwhile, they peaked in total CD4^+^ T cells at one month, which is consistent with the increase in T cell responses observed by ELISpot analysis between two weeks and one month after diagnosis (Fig. 5C, Supplementary Fig. S3B). As in cluster 1, frequencies of CD4^+^AIM^+^ and CD8^+^AIM^+^ T cells tended to be higher compared to other clusters (Fig. 5C, Supplementary Fig. S3A). Also similar to cluster 1, most (63%) participants had six or more symptoms and 25% were hospitalized (Fig. 5D). However, participants in this cluster were mostly younger than 65 years (63%), female (63%) and non-smokers or ex-smokers (75%). Thus, cluster 2 was composed mostly of younger females with more severe disease, but robust antibody responses and T cell responses which may tend towards higher CD4^+^ responses.

The *third cluster* (19% of the subsample) was characterized by antibody responses which were initially high, but declined more sharply than in clusters 1-2. Anti-S-IgG, however, remained detectable in all participants up to the six month follow-up visit. Meanwhile, overall T cell responses initially waned up to three months post-diagnosis, but then increased again for all peptide pools by month six, at which point 92% of participants had detectable N-specific and S2-specific T cells (Fig. 5C, Supplementary Fig. S3A). By flow cytometry, individuals in this cluster tended to have the highest CD8^+^ T cell numbers at two weeks compared to other clusters (Supplementary Fig. S3B). Meanwhile, they showed a decline in CD4^+^ and CD8^+^ T cells and natural killer (NK) cells at one month with a subsequent increase up to six months, which is consistent with the increased T cell responses observed by ELISpot between the three month and six month follow-ups (Fig. 5C). In addition, participants had higher CD19^+^CD27^+^ memory B cells at three and six months compared to the other clusters (Supplementary Fig. S3B). The characteristics of the participants in this cluster were similar to those in the second cluster, but with only 8% requiring hospitalization during acute infection (Fig. 5D). Five of the participants in this cluster had another PCR-test between three and six months after their infection, none of which was positive. Together, cluster 3 appears to represent individuals with more mild disease and more moderate, less-well-sustained antibody responses. The increase in T cell responses between months three and six in these individuals may possibly represent a re-exposure event.

The *fourth cluster* (16% of the subsample) was characterized by the presence of antibodies and a rapid decline of T cells specific to all peptide pools between the two week and one month follow-up visits. By six months post-diagnosis, only 10% had detectable N-specific T cells and none had S2-specific T cells (Fig. 5C, Supplementary Fig. S3A). By flow cytometry, individuals in this cluster had higher total CD4^+^ T cell numbers compared to clusters 2 and 3, but lower compared to clusters 1 and 5 at two weeks and one month, subsequently showing a decline in total CD4^+^ and CD8^+^ T cells up to six months post-diagnosis (Supplementary Fig. S3B). This cluster was characterized by participants who were predominantly male (80%), younger than 65 years (70%), and non- or ex-smokers (100%; Fig. 5D). About 40% reported having six symptoms or more and 40% were hospitalized in the acute phase. Together, cluster 4 appears to be comprised of young males with moderate to severe disease, but who also have less-well-sustained antibody and T cell responses.

The *fifth cluster* (39% of the subsample) was characterized by persistently low and primarily negative antibody responses and variable T cell responses. About half of the participants had detectable M- and N-specific T cells across all assessments. An increase in S1- and S2-specific T cells by three months was noted which was followed by a decline (Fig. 5C, Supplementary Fig. S3A). Despite T cells responses which tended to be lower than for other clusters by ELISpot, these individuals had higher total CD8^+^ T cell numbers compared to all other clusters from one to six months post-diagnosis, as determined by flow cytometric analysis (Supplementary Fig. S3B). They also had high total CD4^+^ T cell numbers compared to other clusters (though not as high as cluster 1 at two weeks to three months), and showed a steady increase in CD4^+^ and CD8^+^ T cells as well as NK cells up to six months (Supplementary Fig. S3B). Most individuals within this cluster were younger than 65 years (80%), female (68%), and had mild disease as reflected by the low reported symptom count with none of the individuals requiring hospitalization (Fig. 5D). That individuals in cluster 5 experience relatively mild disease in the absence of substantial antibody responses suggests compensatory protection by cell-mediated immune responses, possibly including T cells specific to viral proteins not captured by the assays used in this study.

Overall, our findings demonstrate the presence of different joint trajectories of antibody and T cell immune responses after SARS-CoV-2 infection, highlighting the large heterogeneity between individuals. We found that within these clusters, individuals shared similar patterns of immune phenotypes as well as demographic and clinical characteristics.

### Demographic and clinical factors are associated with humoral and cellular immune responses

We then evaluated whether the demographic and clinical factors described within the clusters were associated with antibody responses in the overall study population using adjusted mixed-effects linear regression analyses. Consistent with other reports (*7, 17, 21, 27–30, 48*), we found that older age (≥65 years) (p<0.001), male sex (p=0.011), higher symptom severity defined as having one to five (p<0.001) or more than six COVID-19 symptoms (p<0.001), as well as hospitalization (p=0.006) were statistically significantly associated with higher anti-S-IgG MFI ratios over time (Fig. 6A, Supplementary Table S5). Conversely, being a current smoker was associated with lower responses (p=0.010). Results for anti-S-IgA MFI ratios in the overall population were comparable (Fig. 6B, Supplementary Table S5), and similar trends were identified for anti-N-IgG MFI ratios within the subsample (Fig. 6C). For T cell responses, we found that older age (≥65 years) (p<0.001) and having more than six symptoms (p<0.001) during the acute infection were associated with higher T cell counts over time within the subsample (Fig. 6D, Supplementary Table S6). Sensitivity analyses for antibody or T cell positivity at two weeks and six months after diagnosis using logistic regression models showed comparable trends (Supplementary Tables S7 and S8). Therefore, severity of disease, age and smoking status are associated with the magnitude of immune responses.

**Fig. 6.**
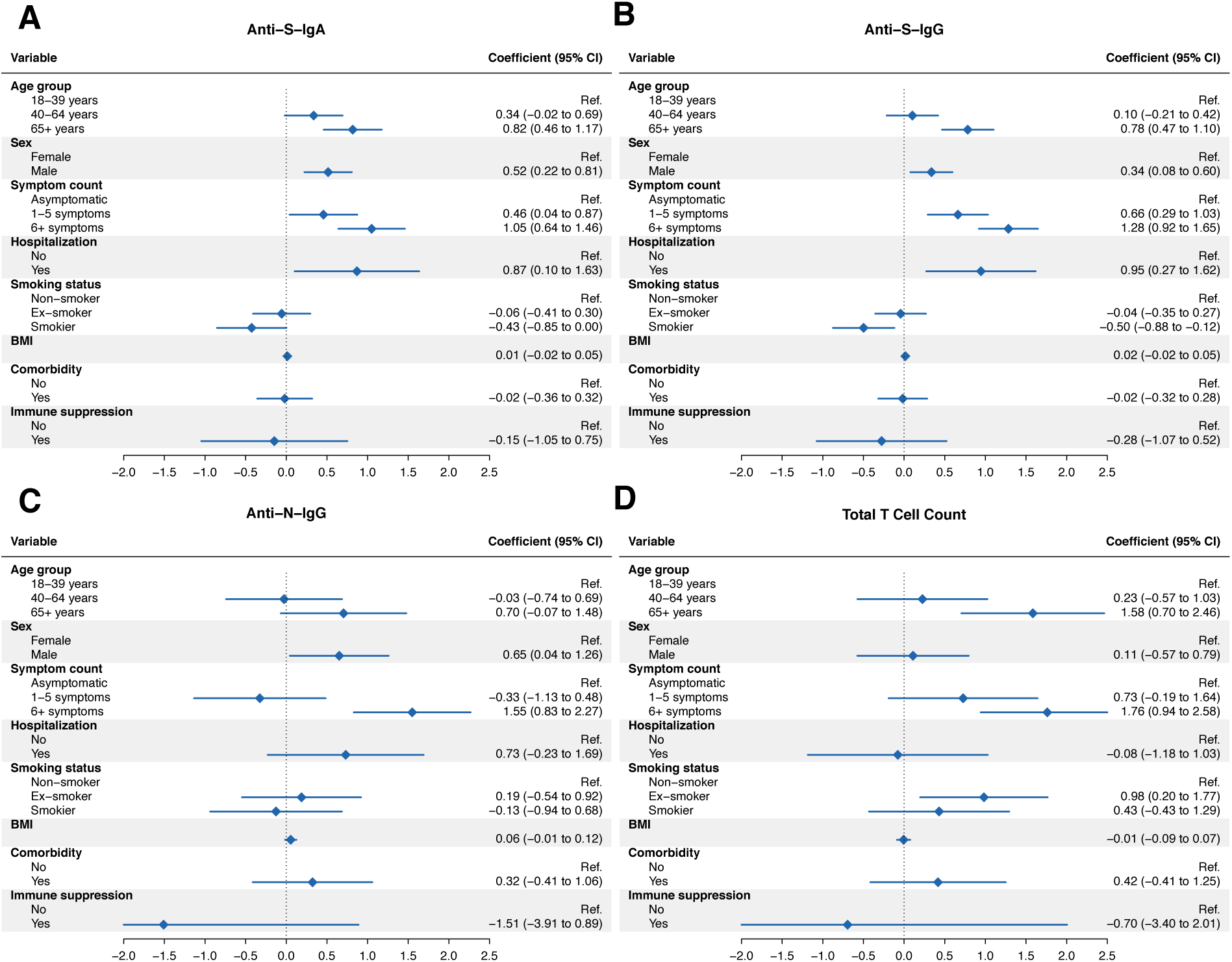
Association analyses of antibody and T cell responses. (**A**) Forest plot demonstrating results from adjusted mixed linear regression analyses evaluating the associations of anti-S-IgA MFI ratios with demographic and clinical factors (age, sex, COVID-19 symptom count, smoking status, body mass index, presence of at least one comorbidity and immunosuppression) in the overall study population (n=431). Model was adjusted for time since diagnosis, age group, sex, and disease severity expressed as symptom count, with a random intercept for each individual. BMI: body mass index. (**B**) Forest plot showing results from adjusted mixed linear regression analyses evaluating the associations of anti-S-IgG MFI ratios with demographic and clinical factors. Model adjustment as for panel A. (**C**) Forest plot showing results from adjusted mixed linear regression analyses evaluating the associations of anti-N-IgG MFI ratios with demographic and clinical factors in the subsample (n=64). Model adjustment as for panel A. (**D**) Forest plot demonstrating results from adjusted mixed linear regression analyses evaluating the associations of overall T cell counts per 1e6 PBMCs with demographic and clinical factors in the subsample (n=64). Model adjustment as for panel A.

## DISCUSSION

Further understanding of the characteristics and trajectories of immune responses after natural infection remains important even with the rollout of vaccines worldwide. Here, we provide a longitudinal evaluation of humoral and cellular immune responses simultaneously, from shortly after, and up to 217 days after, SARS-CoV-2 infection, in a group of individuals infected between August 2020 and January 2021 and covering the full spectrum of clinical manifestation and S-specific antibody responses.

### Distinct kinetics of antibody and T cell responses by subset and specificity

We demonstrate that approximately 85% of individuals infected with SARS-CoV-2 develop S-specific IgA and IgG antibody responses, and that nearly all of these individuals maintain these responses for up to six months of follow-up. Compared to S-specific IgG, however, the proportion of participants with detectable anti-S-IgA decreased markedly over time (from 85% to 70% at six months). Consistent with this, we found the half-life of anti-S-IgA was considerably shorter (71 days, with a range of 42 to 210 days reported by other studies (*2, 8, 18*)) compared to that of anti-S-IgG (145 days, with a range of 36 to 245 days reported by other studies (*2, 8, 11, 33, 49–52*)). This difference, however, is perhaps not surprising as the half-lives of IgA responses, in general, are shorter than those of IgG. Interestingly, we further found that anti-N-IgG responses waned more rapidly than those specific to S, with an estimated half-life of only 86 days. These findings are consistent with those from other studies demonstrating anti-S-IgG responses to be important for sustained protection against SARS-CoV-2. Furthermore, they indicate that both the target of the antibody response as well as the isotype itself appear to influence the duration of humoral immune responses after infection.

In the subsample of individuals selected to cover the full spectrum of clinical disease and S-specific antibody responses, we also found that the majority had detectable, IFN-gamma-producing T cell responses to at least one of the four peptide pools analyzed in this study. Based on this polyclonal T cell response, we estimate a half-life of 145 days, which is consistent with the literature (*2, 8*). Overall T cell positivity in the subsample (nearly 85%) was higher than for overall antibody seropositivity (just over 60%), suggesting that T cell responses are likely able to provide compensatory protection, even in the absence of antibodies. However, as only a limited range of antibody and T cell responses were tested here, it could also be possible that antibody responses to other viral proteins, or of other subtypes, are capable of providing this protection.

Furthermore, the T cell peptide pools for which individuals tested positive were heterogenous. For example, at six months, 50% of individuals were positive for any individual pool despite an overall T cell positivity of approximately 70%, which illustrates the polyclonal nature of the anti-viral T cell response. While M and S1 appeared to be immunodominant in terms of the magnitude of T cell responses, they also decayed more rapidly, with half-lives of 128 and 126 days, compared to N and S2-specific responses with half-lives of 227 and 266 days, respectively. Compared to S1, the S2 domain of the full-length spike protein shares a higher degree of amino acid identity with endemic coronaviruses (*53*). For example, HCoV-HKU1 shares an amino acid identity of 42% with SARS-CoV-2 at the S2 domain of the spike protein compared to 31% at the S1 domain. Therefore, the longer half-life of S2-specific, compared to S1-specific T cells, could potentially be due to intermittent exposure to endemic coronaviruses with similar S2 peptide sequences, resulting in the appearance of a prolonged and more durable T cell response. Overall, our findings suggest that M and S1 responses may initially be more robust, at least in terms of an IFN-gamma-producing response, but that perhaps responses to N and S2 may be more durable.

In evaluating virus-specific CD4^+^ and CD8^+^ T cells by AIM assay we found that frequencies of AIM^+^CD4^+^ or CD8^+^ T cells were similar at two weeks post-infection, but that activated, virus-specific CD4^+^ T cells tended to decline over the six months of follow-up, whereas levels of activated, virus-specific CD8^+^ T cells remained more stable. Furthermore, CD4^+^AIM^+^ T cells were predominantly of TCM phenotype, and this remained consistent throughout the convalescent period, whereas CD8^+^AIM^+^ T cells had a predominantly TEMRA phenotype throughout this period consistent with previous studies (*8, 54, 55*). These findings support the idea that CD4^+^ T cells may play an important role in maintaining the immune response and immunological memory, whereas CD8^+^ T cells may play a more direct role in the anti-viral immune response as highly activated and more terminally-differentiated cytotoxic lymphocytes.

### Concordance of Antibody and T Cell Responses and Immune Response Trajectories

We further evaluated the relationship between antibody and T cell responses within individuals over time, as well as the patterns of these responses within the population. We found a strong, positive correlation between the three antibody subtypes evaluated here. Similarly, T cell responses to M, N, S1, and S2 peptide pools also demonstrated a strong positive correlation, suggesting that, although there may be some variation between distinct subtypes or specificities, both overall antibody and T cell responses tended to behave similarly.

The relationship between antibody and T cell responses, however, was less strong. We observed a weak to moderate positive correlation early after infection, where increased antibody responses tended to predict increased T cell responses to some degree. Meanwhile, correlations became weaker later during follow-up and were weak or no longer present by six months. In assessing the concordance between antibody and T cell responses, we also found that, at two weeks, more than 70% of participants had concordant results (58% both antibody and T cell positive and 13% both antibody and T cell negative). After six months, this dropped to 55-60%. These findings suggest that individuals had differing immune responses following infection (possibly due to differences in viral load or primary site of infection or previous immune history), and that they perhaps retain differing subsets of immune memory components which could be recalled upon reinfection.

Based on this idea, we explored whether there are heterogeneous immune trajectories which individuals tend to follow in response to infection, and which might influence not only their response to infection, but also the immune memory populations which they establish. Using a longitudinal clustering algorithm, we assessed different patterns of immune responses between individuals over time. We identified five distinct joint trajectories of antibody and T cell responses. These trajectories were primarily based on the presence or absence of antibodies and varying T cell responses. Interestingly, we observed distinct clinical characteristics between these clusters. We found that clusters with the most robust immune responses, i.e., clusters 1 and 2, included older participants who had more severe COVID-19 as reflected by hospitalization and the number of reported symptoms. This finding is not surprising as older age and disease severity have been consistently reported to be associated with higher immune responses to SARS-CoV-2 (*7, 17, 21, 27–30*), and other studies primarily conducted in patients with severe COVID-19 or requiring hospitalization observed similar patterns (*4, 16, 21*). These findings were confirmed in association analyses in the overall study population, in which we found that older age, male sex, and higher disease severity were strongly associated with stronger immune responses. In contrast, clusters 4 and 5 appeared to have less robust antibody and T cell responses. Cluster 5, in which participants were distinctly antibody negative, consisted mainly of younger females who reported mild COVID-19 or asymptomatic infection. Some T cell responses were present in half of these participants which, in conjunction with the generally less severe presentation, may point towards compensatory mechanisms of T cell populations in protecting from more severe COVID-19. Remarkably, in cluster 3, we noted a considerable increase in virus-specific T cells, and, to a lesser extent, in anti-S-IgA, after an initial decline up to three months after infection. In combination with the increase in N-specific T cells, this may indicate a possible re-exposure to the virus. Of note, about half of cluster participants underwent SARS-CoV-2 testing between three and six months, which may be indicative of potential exposure events or symptomatic episodes. None, however, tested positive. This again highlights the role of the immune system in protecting from severe disease upon re-exposure, as reinfections may present with milder disease than the primary infection or may even be asymptomatic. Furthermore, this finding demonstrates that reinfections might be more frequent than reported, since many would remain undetected.

### Limitations

Our cohort is one of few population-based and longitudinal studies assessing various components of the immune system in a sample of patients that is representative of the full spectrum of COVID-19. However, some limitations should be considered when interpreting our findings. We used single assays to measure antibodies or T cells in our study. The accuracy and detection levels may differ between tests and thus individuals who are negative in one assay may not be so in another. Nevertheless, the Luminex assay that we used for antibody detection has been extensively validated and was shown to be highly sensitive and specific (*45*). Second, we did not measure the neutralizing capacity of antibody responses. However, other studies have shown that neutralizing capacity correlates strongly with measured levels of binding antibodies (*3, 25, 56*). Third, we limited our T cell analysis to the three dominant antigens for cellular immune responses (S, M and N) (*2, 8, 57*). However, we cannot exclude that in some of the participants, subdominant T cell responses against other viral antigens play important roles, which may have led to an underestimation of the proportion of individuals with T cell responses. Fourth, the cluster analysis bears the limitations that are inherent to the methodology. Using a clustering algorithm that separates the study population into distinct clusters may not be necessarily reflective of clinically meaningful differences. However, we identified distinct clinical and immunological correlates within the different clusters, including various factors (such as the data from flow cytometry) that were not used within the clustering model. Therefore, we consider our results to be relatively robust and leading to a meaningful description of different immune trajectories. Finally, we analyzed antibody and T cell testing results only up to six months, limiting our findings regarding the durability of immune responses. Furthermore, the limited sample size was a pragmatic choice to ensure feasibility of the project, and further immune response patterns may have been observable with additional data. Nonetheless, we believe our study provides a valuable and unique in-depth analysis of joint humoral and cellular immune response trajectories which may lead to further insights on the variability of immune responses to SARS-CoV-2.

### Conclusion

In conclusion, our analyses provide important insights into the different dynamics of antibody and T cell immune responses in individuals covering the entire range from asymptomatic to severe courses of COVID-19. Based on the observed humoral and cellular immune responses, we identify variable immunological trajectories and categorize these into five distinct clusters. While antibody and T cell responses strongly correlate in some individuals, their discordance in other individuals may point towards more complex interactions of the immune system among infected individuals. Overall, our findings indicate that virus-specific T cell responses may be compensatory in the absence of humoral immunity against SARS-CoV-2.

## MATERIALS AND METHODS

### Study Design and Participants

We recruited a population-based, age-stratified, random sample of 431 individuals diagnosed with SARS-CoV-2 infection between 6 August 2020 and 19 January 2021 in the Canton of Zurich, Switzerland. Study participants were identified through the Department of Health of the Canton of Zurich, which records all diagnosed SARS-CoV-2 cases within the Canton through mandatory case reporting. Eligibility criteria were having a polymerase chain reaction (PCR)-confirmed SARS-CoV-2 diagnosis, being aged 18 years or older, residing in the Canton of Zurich, understanding the German language, and being cognitively able to follow the study procedures. We obtained written informed consent from all participants upon study enrollment. The study protocol was approved by the Cantonal Ethics Committee of Zurich (BASEC Registration No. 2020-01739) and prospectively registered (ISRCTN 14990068) (*58*).

We collected peripheral venous blood samples during study visits at two weeks, one month, three months and six months post-diagnosis. Participants additionally provided information regarding acute COVID-19 disease course, severity and symptoms, longer-term health and complications, past medical history, and socio-demographics at the corresponding timepoints through electronic questionnaires.

We selected 64 out of the 431 participants for a detailed characterization of immune responses. This sample was aimed to be representative of the full spectrum of SARS-CoV-2 infection and associated immune responses. Participants in this subsample were selected at random within strata based on clinical characteristics (asymptomatic disease, low and high symptom count, hospitalization) and antibody responses up to one month (negative or low anti-S-IgA or -IgG response, positive or high anti-S-IgA or -IgG response), while ensuring balance across sex and age groups. Based on preliminary assessments and evidence from other studies, we deemed this sample size to be a sensible and pragmatic choice allowing to identify the range and distinct trajectories of immune responses in infected individuals, while ensuring the feasibility of the project.

### Isolation of Plasma and PBMCs

K2 EDTA blood samples collected from participants at each study timepoint were subjected to initial centrifugation to collect plasma, followed by isolation of PBMCs from the remaining cellular fraction by density-gradient centrifugation using Ficoll–Paque (density 1.077g/ml). Plasma aliquots were stored at -20°C prior to IgA and IgG antibody titer analyses. PBMCs were initially frozen at -80°C and transferred to liquid nitrogen prior to use in ELISpot and Activation Induced Marker (AIM) Flow Cytometry assays.

### Analysis of Spike-Specific IgA and IgG and Nucleocapsid-Specific IgG

Frozen plasma samples were thawed and analyzed for levels of Spike (S)-specific IgA and IgG, or Nucleocapsid (N)-specific IgG by Luminex assay as described elsewhere (*45*). In brief, assay beads were prepared by covalent coupling of either the SARS-CoV-2 Spike protein trimer, or N-protein, with MagPlex beads using a Bio-Plex 356 Amine Coupling Kit (Bio-Rad) per manufacturer’s protocol. Protein-coupled beads were diluted and added to each well of Bio-Plex Pro 96-well Flat Bottom Plates (Bio-Rad). Beads were washed with PBS on a magnetic plate washer (MAG2x program) and 50ul of individual serum samples diluted 1:300 in PBS were added to plate wells. A pool of pre-Covid-19 pandemic healthy human sera was used as a negative control (BioWest human serum AB males; VWR). Plates were incubated for 1 hour at room temperature with shaking, washed with PBS and incubated with 50ul of a 1:100 dilution of anti-human IgA-PE (for anti-IgA assay) or IgG-PE (for anti-IgG assay) secondary antibody (OneLambda ThermoFisher) at room temperature for an additional 45 minutes with shaking. After incubation, samples were washed with PBS and resuspended in reading buffer and read on a Luminex FLEXMAP 3D plate reader (ThermoFisher) to obtain a Mean Fluorescence Intensity (MFI) value for each sample. The MFI value for each serum sample was divided by the mean value of the negative control samples to yield an MFI ratio. Based on negative control samples and samples from PCR-positive donors, cut-off thresholds for MFI ratios used to determine seropositivity were 6.5 for IgA and 6.0 for IgG. The lower limit of measured MFI ratios was restricted to 1, representing equivalent fluorescence intensity compared to negative control samples. For anti-S-IgG MFI ratios, an approximate conversion to BAU/ml is available based on a cross-validation with the Roche Elecsys Anti-SARS-CoV-2 S immunoassay, for which a reference table is provided (Supplementary Table S9).

### ELISpot Assay

T cell responses were assessed by ELISpot assay using the Human IFN-gamma ELISpot Assay kit (R&D Systems) following the manufacturer’s instructions. For the assay, cryopreserved PBMCs were thawed and plated at 5e5 cells per well. Cells were stimulated 20h at 37°C with overlapping 15mer peptide pools spanning the entire M and N proteins or the S1 domain of the spike protein or a mix of the predicted immunodominant peptides from the spike protein containing the majority of the S2 domain (M, N, S1 and S PepTivator peptide pools, respectively; Miltenyi Biotec) at a concentration of 1ug/ml per individual peptide. As negative controls, cells were incubated without peptide. As positive controls, 2.5e5 cells per well were stimulated with anti-CD3 antibody (OKT3; Miltenyi Biotec). Spots were counted using an AID iSpot Reader System with EliSpot 7.0 software (AID). Two times the number of spots in unstimulated negative control wells were subtracted from the values of each test well and results were normalized to the number of spots in anti-CD3 antibody-stimulated wells from the same individual and timepoint and presented as spot-forming units (SFU) per 1e6 CD3+ cells. Results were excluded if positive control wells were negative.

### Flow Cytometry and Activation-Induced Marker (AIM) Assay

Cryopreserved PBMCs were thawed in RPMI-1640 (Gibco, Thermo Fisher Scientific) supplemented with 5% human AB-serum (BioConcept) and 25U/ml benzonase (Sigma), and plated in 96-UWell plates (Sarstedt) at a concentration of up to 1e6 cells per well in RPMI-1640 medium (Gibco, Thermo Fisher Scientific) supplemented with 10% human AB-serum (BioConcept) and 1% Penicillin-Streptomycin (Thermo Fisher). SARS-CoV-2 PepTivator peptide pools M, N, S1 and S (Miltenyi Biotec) were dissolved per manufacturer’s instructions in sterile water and combined into a single mega pool. PBMCs were cultured for 24h in a humidified incubator at 37°C and 5% CO_2_ in the presence of either the SARS-CoV-2 mega pool at 0.6nmol (appr. 1μg) of each peptide/ml, Phytohemagglutinin-L at 5μg/ml (Merck Millipore, positive control) or culture medium (unstimulated condition). Peptide- and unstimulated samples were run in duplicate whenever possible and longitudinal samples from individual participants were included in the same assay. After 24 hours, cells were washed in staining buffer (PBS, 0.02% NaN_3_, 2mM EDTA, 1% bovine serum albumin), blocked for 10 minutes with Human TruStain FcX (Biolegend) on ice and stained for 30 minutes at 4°C with the following antibodies in buffer supplemented with Super Bright Complete Staining Buffer (eBioscience): BUV395 anti-CD45RA (Clone: HI100, BD Bioscience, RRID:AB_2740037), BUV496 anti-CD8 (Clone: RPA-T8, BD Bioscience, RRID:AB_2870223), BUV563 anti-CD56 (Clone: NCAM16.2, BD Bioscience, RRID:AB_2870213), BUV661 anti-CD14 (Clone: M5E2, BD Bioscience, RRID:AB_2871011), BUV737 anti-CD16 (Clone: 3G8, BD Bioscience, RRID:AB_2869578), BUV805 anti-CD19 (Clone: SJ25C1, BD Bioscience, RRID:AB_2873553), BV421 anti-CD27 (Clone: O323, Biolegend, RRID:AB_11150782), BV510 anti-CD4 (Clone: OKt4, Biolegend, RRID:AB_2561866), BV650 anti-CD38 (Clone: HB-7, Biolegend, RRID:AB_2566233), BV786 anti-CD3 (Clone: OKt3, Biolegend, RRID:AB_2563507), PE anti-IgD (Clone: IA6-2, Biolegend, RRID:AB_10553900), PE/Dazzle594 anti-CCR7 (Clone: G043H7, Biolegend, RRID:AB_2563641), FITC anti-HLA-DR (Clone: L243, Biolegend, RRID:AB_314682), PE-Cy7 anti-CD137 (Clone: 4B4-1, Biolegend, RRID:AB_2207741), BB700 anti-CD134/OX40 (Clone: ACT35, BD Bioscience, RRID:AB_2743451), APC anti-CD69 (Clone: FN50, Biolegend, RRID:AB_314845). Zombie NIR Fixable Viability Dye (Biolegend) was used to exclude dead cells. After washing, samples were fixed with 1% PFA and acquired on Cytek Aurora 5L spectral flow cytometer (Cytek). Data was analyzed using Cytek SpectroFlo (version 3.0.1) and FlowJo software (version 10, TreeStar Inc). Samples were excluded if the percentage of live leukocytes was below 25%. SARS-CoV-2 antigen-specific T cells were measured as AIM^+^ (CD134^+^CD137^+^) CD4^+^ T and (CD69^+^CD137^+^) CD8^+^ T cells. Unspecific activation in unstimulated controls was subtracted and negative values were set to zero.

### Statistical Analyses

We summarized population characteristics descriptively and report frequencies and percentage or median and interquartile range, as applicable. We report age-stratified summary statistics for the frequency of antibody and T cell responses and calculated 95% Wilson confidence intervals to estimate the associated uncertainty. We excluded any data measured after receipt of COVID-19 vaccination (first dose; n=2 participants at three months and n=78 participants at six months) or diagnosed reinfection (based on self-reported positive PCR or rapid antigen test; n=3 participants at six months) from all analyses. All data were transformed using natural logarithms for all comparative and associational analyses due to non-normal distributions and ratio properties of the Ig MFI ratios (zero values being replaced by half of the lowest non-zero value). Results are visualized using a log10-transformation in figures.

To estimate antibody decay times, we first determined the maximum response timepoint for each individual. We excluded data from individuals that were never tested positive for the respective antibody (i.e., anti-S-IgA or -IgG, or anti-N-IgG). We then restricted the data to the maximum and all subsequent timepoints and rescaled the time axis to start with the maximum concentration, in order to restrict and align the data for the descending slope of antibody decay (in line with previous studies (*59, 60*)). For T cells, we used the data as measured, as peak T cell expansion typically occurs in the first week after infection (*61*). We then fitted uni- and multivariable mixed-effects linear decay models on the natural logarithm-transformed data using random intercepts for individuals. We then calculated the half-life in days using the formula *λ* = *ln*(0.5) / *β*, where β is the model-derived intercept (and associated uncertainty bounds). We used mixed-model-based parametric bootstrap for the visualization of confidence bounds.

For assessing the correlation of antibody and T cell test results, we calculated Spearman correlation coefficients for all combinations of antibody subtypes and epitope-specific T cells. Furthermore, we assessed concordance by calculating the proportion of participants testing positive or negative for antibody subtypes and overall T cells. And last, we further assessed agreement by calculating unweighted Cohen’s Kappa values for test positivity for all possible combinations of antibody subtypes and virus-specific T cells.

We assessed the association between demographic and clinical factors and IgA and IgG MFI ratios up to six months after diagnosis using uni- and multivariable mixed-effects linear regression models. Model selection was based on prior knowledge and the Akaike and Bayesian Information Criteria (AIC/BIC), with a difference of 2 points considered relevant. Age, sex, and time since diagnosis were defined as a priori variables based on findings from previous studies. We conducted sensitivity analyses for antibody positivity at two weeks and six months after diagnosis using uni- and multivariable logistic regression analyses.

To identify clusters of individuals with similar immune response trajectories over the four assessment timepoints, we used the *KmL3D k*-means clustering method which allows the joint evolution of multiple variables with repeated measures (*46, 47*). The algorithm requires predefining the number of clusters. We chose four to six clusters based on a priori knowledge on immune response patterns as well as exploratory analyses of the data. We specified 100 runs for each *k* clusters (i.e., 300 times in total) and specified Euclidean distance with Gower adjustment to estimate similarity between the trajectories. The selection of the final number of clusters was based on maximizing the Calinski and Harabatz quality criterion (*46, 47*) as well as the expected patterns in the data. Implementing the *KmL3D* algorithm requires that data for all variables included in the analysis are available for all participants. Hence, missing data were imputed by applying linear extrapolation with added variation (“Copy Mean” function (*46, 47*)). We plotted the mean antibody and T cell responses for each cluster to explore differences in respective immune response trajectories. Finally, we descriptively compared demographic and clinical features of individuals in clusters to identify specific factors associated with each trajectory.

All analyses were performed using *R* (v4.1.1) (*62*), using the *Hmisc* (v4.5-0), *lme4* (v1.1-27.1), *lmerTest* (v3.1-3) and *KmL3D* (v2.4.2) packages, and results were visualized using the *ggplot2* (v3.3.5), *ggpubr* (v0.4.0) and *pheatmap* (v1.0.12) packages.

## Data Availability

All data are available in the main text or the supplementary materials.

## Supplementary Materials

Fig. S1. Overall antibody and T cell positivity across timepoints and estimation of T cell decay kinetics.

Fig. S2. Concordance of antibody and T cell positivity over time.

Fig. S3. Antibody responses, cellular subsets and AIM+ T cells within clusters.

Table S1. Study population characteristics.

Table S2. Anti-S-IgA and -IgG antibody responses in the overall study population over time.

Table S3. Antibody and T cell responses in the subsample over time.

Table S4. Sensitivity analysis regarding anti-S-IgA and -IgG antibody responses, weighted by age group.

Table S5. Association between demographic and clinical factors and antibody responses over time.

Table S6. Association between demographic and clinical factors and T cell responses over time.

Table S7. Association between demographic and clinical factors and anti-S-IgG antibody positivity at two weeks and six months.

Table S8. Association between demographic and clinical factors and overall T cell positivity at two weeks and six months.

Table S9. Reference table for converting anti-S-IgG MFI ratios to BAU/ml (based on Roche Elecsys Anti-SARS-CoV-2 S immunoassay).

## Acknowledgments

The authors would like to thank the study administration team and the study participants for their dedicated contribution to this research project.

## Funding

This study is part of Corona Immunitas research network, coordinated by the Swiss School of Public Health (SSPH+), and funded by fundraising of SSPH+ that includes funds of the Swiss Federal Office of Public Health and private funders (ethical guidelines for funding stated by SSPH+ will be respected), by funds of the Cantons of Switzerland (Vaud, Zurich, and Basel) and by institutional funds of the Universities. Additional funding, specific to this study was received from the Department of Health of the Canton of Zurich, the University of Zurich Foundation, and the Swiss Federal Office of Public Health. The funding bodies had no influence on the study design or the conduct, analysis, or interpretation of the study findings. TB received funding from the European Union’s Horizon 2020 research and innovation programme under the Marie Skłodowska-Curie grant agreement No 801076, through the SSPH+ Global PhD Fellowship Programme in Public Health Sciences (GlobalP3HS) of the SSPH+. HA received an Swiss National Science Foundation (SNSF) Early Postdoc.Mobility Fellowship.

## Author contributions

Conceptualization: DM, KDZ, TB, CM, MAP, JSF

Methodology: DM, KDZ, TB, NC, CP, MP, CF, GP, CM, MAP, JSF

Investigation: DM, KDZ, TB, NC, CP, MP, CF, GP, CRK

Visualization: DM, KDZ, TB, NC

Funding acquisition: MAP, JSF

Project administration: DM, TB, HEA, AD, MAP

Supervision: CM, MAP, JSF

Writing – original draft: DM, KDZ, TB

Writing – review & editing: DM, KDZ, TB, NC, DLC, HEA, AD, CP, MP, CF, GP, CRK, CM, MAP, JSF

## Competing interests

The authors declare that they have no competing interests.

## Data and materials availability

All data are available in the main text or the supplementary materials.

## Notes

### Competing Interest Statement

The authors have declared no competing interest.

### Author Declarations

The responsible Ethics Committee of the Canton of Zurich, Switzerland, gave ethical approval for this work(BASEC Registration No. 2020-01739).

