## Supplementary Material for "Heterogenous Cellular and Humoral Immune Trajectories after SARS-CoV-2 Infection: Compensatory Responses in a Population-Based Cohort"

**Supplementary Figures**

Fig. S1. Overall antibody and T cell positivity across timepoints and estimation of T cell decay kinetics.

Fig. S2. Concordance of antibody and T cell positivity over time.

Fig. S3. Antibody responses, cellular subsets and AIM+ T cells within clusters.

**Supplementary Tables**

Table S1. Study population characteristics.

Table S2. Anti-S-IgA and -IgG antibody responses in the overall study population over time.

Table S3. Antibody and T cell responses in the subsample over time.

Table S4. Sensitivity analysis regarding anti-S-IgA and -IgG antibody responses, weighted by age group.

Table S5. Association between demographic and clinical factors and antibody responses over time.

Table S6. Association between demographic and clinical factors and T cell responses over time.

Table S7. Association between demographic and clinical factors and anti-S-IgG antibody positivity at two weeks and six months.

Table S8. Association between demographic and clinical factors and overall T cell positivity at two weeks and six months.

Table S9. Reference table for converting anti-S-IgG MFI ratios to BAU/ml (based on Roche Elecsys Anti-SARS-CoV-2 S immunoassay).

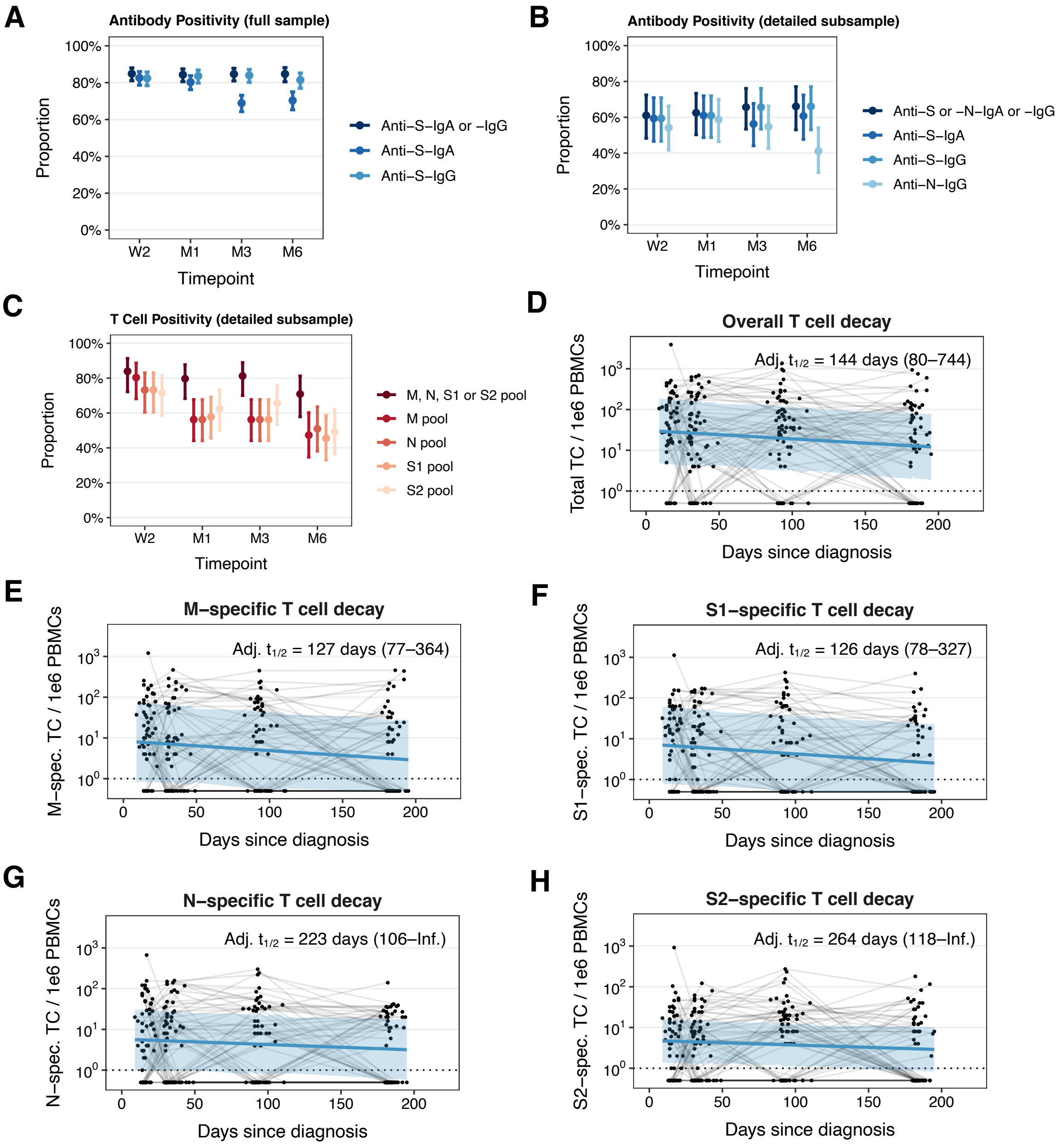


**Fig. S1**. **Overall antibody and T cell positivity across timepoints and estimation of T cell decay kinetics.** (**A**) Seropositivity for anti-S-IgA and anti-S-IgG antibodies across timepoints in the overall study population (n=431). W2: two weeks, M1: one month, M3: three months, M6: six months after diagnosis. (**B**) Seropositivity for anti-S-IgA, anti-S-IgG and anti-N-IgG antibodies across timepoints in the subsample selected for detailed examination of immune responses (n=64). (**C**) Positivity for virus-specific T cells (measured by ELISpot for M, N, S1 and S2 antigen pools) across timepoints in the subsample (n=64). (**D**) Decay estimation for total T cell count based on mixed linear regression model. Adj. $t_{1/2}$: half-life based on model adjusted for time from diagnosis to maximum MFI ratio, age group, sex and symptom count, using a random intercept for each individual in the study. (**E**) Decay estimation for M-specific T cells based on mixed linear regression model. (**F**) Decay estimation for N-specific T cells based on mixed linear regression model. (**G**) Decay estimation for S1-specific T cells based on mixed linear regression model. (**H**) Decay estimation for S2-specific T cells based on mixed linear regression model.

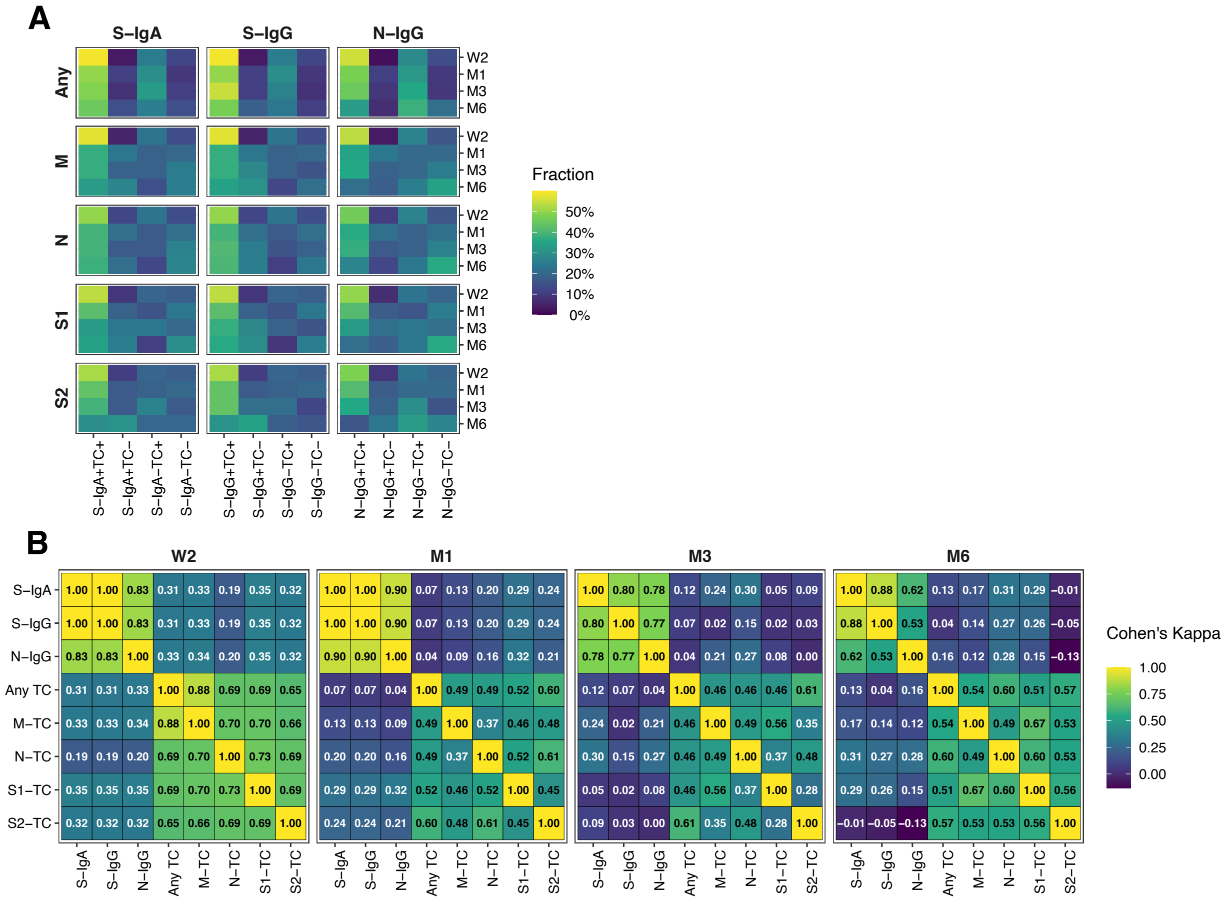


**Fig. S2. Concordance of antibody and T cell positivity over time.** (**A**) Percent concordance of positivity in antibody subtypes compared to positivity of overall (any) and virus-specific T cells (based on M, N, S1 and S2 epitopes). W2: two weeks, M1: one month, M3: three months, M6: six months after diagnosis. (**B**) Concordance of antibody subtype and virus-specific T cell test results calculated based on Cohen's Kappa.

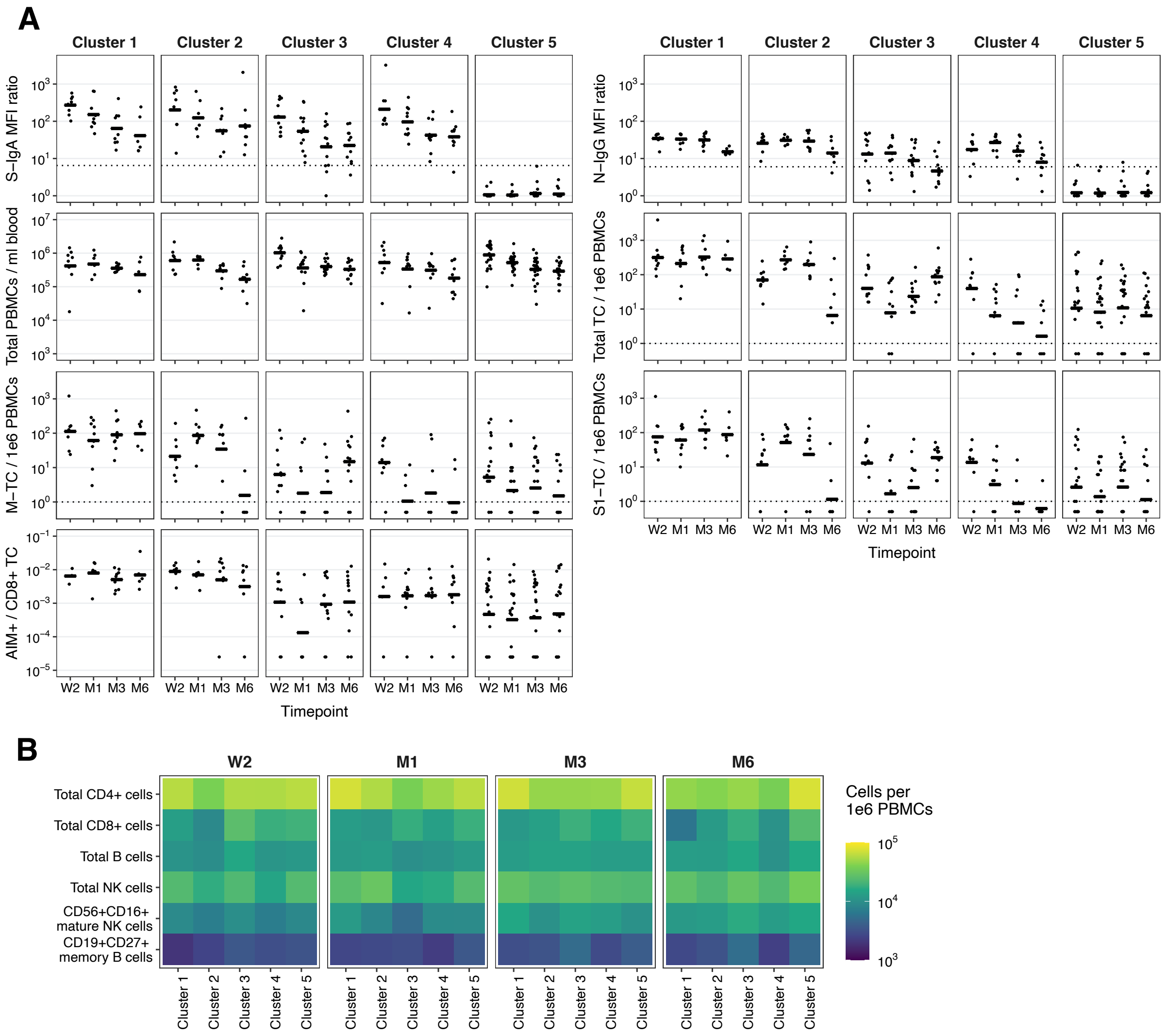


**Fig. S3**. **Antibody responses, cellular subsets and AIM+ T cells within clusters.** (**A**) Dot plot based on antibody testing, ELISpot and flow cytometric analyses in the detailed subsample (n=64), stratified by cluster. Plots demonstrate results for anti-S-IgA and anti-N-IgG MFI ratios, total PBMCs / ml blood, total T cells / 1e6 PBMCs, N-specific T cells/1e6 PBMCs, S1-specific T cells/1e6 PBMCs, and frequency of AIM$^{+}$ of CD8$^{+}$ T cells. Horizontal lines represent mean at timepoint. TC: T cells, MFI: mean fluorescence intensity, W2: two weeks, M1: one month, M3: three months, M6: six months after diagnosis. (**B**) Heatmap based on flow cytometric analyses stratified by cluster. Heatmap demonstrates the total number of $CD4^+$ T cells, $CD8^+$ T cells, B cells, NK cells, $CD56^+CD16^+$ B cells, and $CD19^+CD27^+$ NK cells per 1e6 PBMCs. NK: natural killer.

**Table S1**. **Study population characteristics.** The table displays the characteristics of the overall study sample (n=431) and of the subsample (n=64) selected for detailed immune analyses aimed to cover the full spectrum of SARS-CoV-2 infection and associated immune responses. Study participants were recruited between 06 August 2020 and 26 January 2021. CHF: Swiss Francs, IQR: Interquartile Range, SD: Standard Deviation.

| | **Overall Study Population (N=431)** | **Subsample Participants (N=64)** | **Participants not in the Subsample (N=367)** |
|---|---|---|---|
| **Age** | | | |
| Mean (SD) | 51.7 (18.3) | 52.1 (18.9) | 51.6 (18.3) |
| Median (IQR) | 52.0 (35.0 to 68.0) | 53.5 (33.5 to 68.0) | 52.0 (35.0 to 68.0) |
| Range | 18 to 88 | 18 to 87 | 18 to 88 |
| **Age group** | | | |
| 18-39 years | 135 (31.3%) | 21 (32.8%) | 114 (31.1%) |
| 40-64 years | 144 (33.4%) | 21 (32.8%) | 123 (33.5%) |
| 65+ years | 152 (35.3%) | 22 (34.4%) | 130 (35.4%) |
| **Sex** | | | |
| female | 212 (49.2%) | 36 (56.2%) | 176 (48.0%) |
| male | 219 (50.8%) | 28 (43.8%) | 191 (52.0%) |
| **Symptom count at diagnosis** | | | |
| Asymptomatic | 76 (17.6%) | 20 (31.2%) | 56 (15.3%) |
| 1-5 symptoms | 163 (37.8%) | 17 (26.6%) | 146 (39.8%) |
| ≥6 symptoms | 192 (44.5%) | 27 (42.2%) | 165 (45.0%) |
| **Symptom count at diagnosis** | | | |
| Mean (SD) | 5.1 (3.8) | 5.2 (5.4) | 5.1 (3.5) |

| | | | |
|---|---|---|---|
| Median (IQR) | 5.0 (2.0 to 7.0) | 4.0 (0.0 to 8.5) | 5.0 (3.0 to 7.0) |
| Range | 0 to 20 | 0 to 20 | 0 to 16 |
| **Self-reported symptom severity at diagnosis** | | | |
| None | 74 (17.3%) | 20 (31.7%) | 54 (14.8%) |
| Mild to moderate | 281 (65.7%) | 30 (47.6%) | 251 (68.8%) |
| Severe to very severe | 73 (17.1%) | 13 (20.6%) | 60 (16.4%) |
| Missing | 3 (0.7%) | 1 (1.6%) | 2 (0.5%) |
| **Hospitalisation within first 2 weeks** | | | |
| No | 413 (95.8%) | 53 (82.8%) | 360 (98.1%) |
| Yes | 18 (4.2%) | 11 (17.2%) | 7 (1.9%) |
| **ICU admission within first 2 weeks** | | | |
| No | 429 (99.5%) | 63 (98.4%) | 366 (99.7%) |
| Yes | 2 (0.5%) | 1 (1.6%) | 1 (0.3%) |
| **Vaccinated at 6 months follow-up** | | | |
| No | 351 (81.4%) | 57 (89.1%) | 294 (80.1%) |
| Yes | 80 (18.6%) | 7 (10.9%) | 73 (19.9%) |
| **Reinfected at 6 months follow-up** | | | |
| No | 428 (99.3%) | 64 (100.0%) | 364 (99.2%) |
| Yes | 3 (0.7%) | 0 (0.0%) | 3 (0.8%) |
| **Smoking status** | | | |
| Non-smoker | 261 (61.3%) | 29 (46.0%) | 232 (63.9%) |
| Ex-smoker | 106 (24.9%) | 20 (31.7%) | 86 (23.7%) |
| Smoker | 59 (13.8%) | 14 (22.2%) | 45 (12.4%) |
| Missing | 5 (1.2%) | 1 (1.6%) | 4 (1.1%) |
| **Body mass index** | | | |
| Mean (SD) | 24.5 (4.1) | 25.4 (4.5) | 24.3 (4.0) |
| Missing | 5 (1.2%) | 1 (1.6%) | 4 (1.1%) |

| | | | |
|---|---|---|---|
| **Comorbidities** | | | |
| No | 300 (69.6%) | 44 (68.8%) | 256 (69.8%) |
| Yes | 131 (30.4%) | 20 (31.2%) | 111 (30.2%) |
| **Immune suppression** | | | |
| No | 410 (97.4%) | 61 (98.4%) | 349 (97.2%) |
| Yes | 11 (2.6%) | 1 (1.6%) | 10 (2.8%) |
| Missing | 10 (2.3%) | 2 (3.1%) | 8 (2.2%) |
| **Income** | | | |
| <6'000 CHF | 139 (33.7%) | 20 (32.3%) | 119 (33.9%) |
| 6'000 - 12'000 CHF | 182 (44.1%) | 33 (53.2%) | 149 (42.5%) |
| >12'000 CHF | 92 (22.3%) | 9 (14.5%) | 83 (23.6%) |
| Missing | 18 (4.2%) | 2 (3.1%) | 16 (4.4%) |
| **Employment** | | | |
| Employed | 258 (59.9%) | 37 (57.8%) | 221 (60.2%) |
| Student | 22 (5.1%) | 4 (6.2%) | 18 (4.9%) |
| Retired | 124 (28.8%) | 18 (28.1%) | 106 (28.9%) |
| Unemployed or other | 27 (6.3%) | 5 (7.8%) | 22 (6.0%) |
| **Education** | | | |
| None or mandatory school | 18 (4.2%) | 4 (6.2%) | 14 (3.8%) |
| Vocational training or specialized baccalaureate | 179 (41.5%) | 27 (42.2%) | 152 (41.4%) |
| Higher technical school or college | 113 (26.2%) | 14 (21.9%) | 99 (27.0%) |
| University | 121 (28.1%) | 19 (29.7%) | 102 (27.8%) |
| **Nationality** | | | |
| Swiss | 376 (87.2%) | 53 (82.8%) | 323 (88.0%) |
| Non-Swiss | 55 (12.8%) | 11 (17.2%) | 44 (12.0%) |

**Table S2**. **Anti-S-IgA and -IgG antibody responses in the overall study population over time.** The table shows detailed antibody test results for anti-S-IgA, -IgG or combined anti-S antibody responses in the full study population (n=431) at two weeks, one month, three months and six months after diagnosis of SARS-CoV-2 infection. Data measured after vaccination or known reinfection were removed to allow analysis of immune trajectories. Anti-S-IgG MFI ratios were converted into BAU/ml (based on Roche Elecsys Anti-SARS-CoV-2 IgG) based on a formula derived from cross-validations. IQR: Interquartile Range, SD: Standard Deviation.

| | **Overall Study Population** | | | |
|---|---|---|---|---|
| | **2 Weeks (N=404)** | **1 Month (N=421)** | **3 Months (N=418)** | **6 Months (N=334)** |
| **Anti-S IgA** | | | | |
| negative | 70 (17.3%) | 83 (19.7%) | 130 (31.1%) | 99 (29.6%) |
| positive | 334 (82.7%) | 338 (80.3%) | 288 (68.9%) | 235 (70.4%) |
| **Anti-S IgA MFI ratio** | | | | |
| Mean (SD) | 137.4 (242.1) | 75.0 (132.1) | 37.9 (90.1) | 39.4 (149.3) |
| Median (IQR) | 65.2 (18.8 to 155.2) | 36.0 (9.9 to 77.7) | 12.7 (5.1 to 35.8) | 12.8 (4.8 to 31.1) |
| Range | 0 to 3216 | 0 to 1179 | 0 to 1204 | 0 to 2058 |
| **Anti-S IgG** | | | | |
| negative | 71 (17.6%) | 69 (16.4%) | 67 (16.0%) | 62 (18.6%) |
| positive | 333 (82.4%) | 352 (83.6%) | 351 (84.0%) | 272 (81.4%) |
| **Anti-S IgG MFI ratio** | | | | |
| Mean (SD) | 38.7 (34.0) | 43.9 (37.8) | 41.4 (37.6) | 27.6 (27.4) |
| Median (IQR) | 32.2 (12.0 to 57.9) | 37.1 (16.1 to 61.8) | 30.4 (13.6 to 60.5) | 21.1 (8.2 to 36.4) |

| | | | | |
|---|---|---|---|---|
| Range | 0 to 158 | 0 to 185 | 0 to 187 | 0 to 156 |
| **Anti-S IgG MFI ratio (converted to BAU/ml for Roche anti-S IgG)** | | | | |
| Mean (SD) | 689.2 (1044.4) | 873.1 (1352.3) | 812.1 (1316.2) | 394.7 (791.7) |
| Median (IQR) | 277.0 (41.2 to 875.6) | 365.2 (73.1 to 995.9) | 248.5 (53.1 to 954.2) | 121.8 (21.2 to 351.4) |
| Range | 1 to 6452 | 1 to 8787 | 1 to 8978 | 1 to 6234 |
| **Anti-S IgA or IgG** | | | | |
| negative | 61 (15.1%) | 66 (15.7%) | 64 (15.3%) | 51 (15.3%) |
| positive | 343 (84.9%) | 355 (84.3%) | 354 (84.7%) | 283 (84.7%) |

**Table S3**. **Antibody and T cell responses in subsample over time.** The table shows the detailed results for anti-S-IgA or -IgG and anti-N-IgG antibodies, as well as M, N, S and S1 pool-specific T cell responses in the subsample selected for detailed analyses (n=64) at two weeks, one month, three months and six months after diagnosis of SARS-CoV-2 infection. Antibody responses were quantified as mean fluorescence intensity (MFI) ratios compared to pre-pandemic seronegative control samples. Anti-S-IgG MFI ratios were converted into BAU/ml (based on Roche Elecsys Anti-SARS-CoV-2 IgG) based on a formula derived from cross-validations. IQR: Interquartile Range, SD: Standard Deviation.

| | **Subsample Participants** | | | |
|---|---|---|---|---|
| | **2 Weeks (N=59)** | **1 Month (N=64)** | **3 Months (N=64)** | **6 Months (N=56)** |
| **Anti-S IgA** | | | | |
| negative | 24 (40.7%) | 25 (39.1%) | 28 (43.8%) | 22 (39.3%) |
| positive | 35 (59.3%) | 39 (60.9%) | 36 (56.2%) | 34 (60.7%) |
| **Anti-S IgA MFI ratio** | | | | |
| Mean (SD) | 201.6 (446.3) | 97.3 (156.0) | 43.7 (70.2) | 70.3 (275.3) |
| Median (IQR) | 82.1 (0.6 to 274.9) | 41.0 (0.5 to 120.8) | 12.4 (0.5 to 49.8) | 16.2 (0.7 to 47.5) |
| Range | 0 to 3216 | 0 to 646 | 0 to 407 | 0 to 2058 |
| **Anti-S IgG** | | | | |
| negative | 24 (40.7%) | 25 (39.1%) | 22 (34.4%) | 19 (33.9%) |
| positive | 35 (59.3%) | 39 (60.9%) | 42 (65.6%) | 37 (66.1%) |
| **Anti-S IgG MFI ratio** | | | | |
| Mean (SD) | 37.0 (39.6) | 40.8 (43.8) | 45.0 (49.4) | 31.4 (33.4) |

| | | | | |
|---|---|---|---|---|
| Median (IQR) | 24.0 (0.5 to 73.5) | 33.2 (0.5 to 72.6) | 29.6 (0.5 to 64.8) | 19.2 (0.5 to 47.0) |
| Range | 0 to 129 | 0 to 159 | 0 to 187 | 0 to 120 |
| **Anti-S IgG MFI ratio (converted to BAU/ml for Roche anti-S IgG)** | | | | |
| Mean (SD) | 756.9 (1079.5) | 921.6 (1476.5) | 1145.1 (1866.6) | 542.3 (844.7) |
| Median (IQR) | 156.8 (1.0 to 1402.7) | 294.9 (1.0 to 1369.5) | 236.0 (1.0 to 1092.6) | 102.7 (1.0 to 580.9) |
| Range | 1 to 4297 | 1 to 6525 | 1 to 8978 | 1 to 3746 |
| **Anti-N IgG** | | | | |
| negative | 27 (45.8%) | 26 (41.3%) | 29 (45.3%) | 33 (58.9%) |
| positive | 32 (54.2%) | 37 (58.7%) | 35 (54.7%) | 23 (41.1%) |
| *Missing* | 0 (0%) | 1 (1.6%) | 0 (0%) | 0 (0%) |
| **Anti-N IgG MFI ratio** | | | | |
| Mean (SD) | 16.5 (17.3) | 17.1 (16.6) | 15.0 (16.8) | 7.9 (8.9) |
| Median (IQR) | 8.4 (0.4 to 31.4) | 13.6 (0.6 to 31.4) | 8.2 (0.7 to 23.8) | 4.3 (0.6 to 12.9) |
| Range | 0 to 48 | 0 to 46 | 0 to 57 | 0 to 39 |
| *Missing* | 0 (0%) | 1 (1.6%) | 0 (0%) | 0 (0%) |
| **Anti-S IgA or IgG** | | | | |
| negative | 24 (40.7%) | 25 (39.1%) | 22 (34.4%) | 19 (33.9%) |
| positive | 35 (59.3%) | 39 (60.9%) | 42 (65.6%) | 37 (66.1%) |
| **Anti-S or Anti-N IgG** | | | | |
| negative | 23 (39.0%) | 24 (37.5%) | 22 (34.4%) | 19 (33.9%) |
| positive | 36 (61.0%) | 40 (62.5%) | 42 (65.6%) | 37 (66.1%) |
| **Anti-S IgA or IgG or Anti-N IgG** | | | | |
| negative | 23 (39.0%) | 24 (37.5%) | 22 (34.4%) | 19 (33.9%) |
| positive | 36 (61.0%) | 40 (62.5%) | 42 (65.6%) | 37 (66.1%) |

| | | | | |
|---|---|---|---|---|
| **Anti-M T cells** | | | | |
| negative | 11 (19.6%) | 28 (43.8%) | 28 (43.8%) | 29 (52.7%) |
| positive | 45 (80.4%) | 36 (56.2%) | 36 (56.2%) | 26 (47.3%) |
| *Missing* | 3 (5.1%) | 0 (0%) | 0 (0%) | 1 (1.8%) |
| **Anti-N T cells** | | | | |
| negative | 15 (26.8%) | 28 (43.8%) | 28 (43.8%) | 27 (49.1%) |
| positive | 41 (73.2%) | 36 (56.2%) | 36 (56.2%) | 28 (50.9%) |
| *Missing* | 3 (5.1%) | 0 (0%) | 0 (0%) | 1 (1.8%) |
| **Anti-S1 T cells** | | | | |
| negative | 15 (26.8%) | 27 (42.2%) | 28 (43.8%) | 30 (54.5%) |
| positive | 41 (73.2%) | 37 (57.8%) | 36 (56.2%) | 25 (45.5%) |
| *Missing* | 3 (5.1%) | 0 (0%) | 0 (0%) | 1 (1.8%) |
| **Anti-S T cells** | | | | |
| negative | 16 (28.6%) | 24 (37.5%) | 22 (34.4%) | 28 (50.9%) |
| positive | 40 (71.4%) | 40 (62.5%) | 42 (65.6%) | 27 (49.1%) |
| *Missing* | 3 (5.1%) | 0 (0%) | 0 (0%) | 1 (1.8%) |
| **Anti-M, -N, -S1, or -S T cells** | | | | |
| negative | 9 (16.1%) | 13 (20.3%) | 12 (18.8%) | 16 (29.1%) |
| positive | 47 (83.9%) | 51 (79.7%) | 52 (81.2%) | 39 (70.9%) |
| *Missing* | 3 (5.1%) | 0 (0%) | 0 (0%) | 1 (1.8%) |

**Table S4. Sensitivity analysis regarding anti-S-IgA and -IgG antibody responses, weighted by age group.** The table presents assay positivity for anti-S-IgA, -IgG and either -IgA or -IgG, based on the primary analysis (unweighted) and based on a sensitivity analysis applying weighting by age groups based on the total number of identified cases during the study timeframe.

| Assay positivity | unweighted | | | |
|---|---|---|---|---|
| | W2 (N=404) | M1 (N=421) | M3 (N=418) | M6 (N=334) |
| **Anti-S IgA** | 82.7% | 80.3% | 68.9% | 70.4% |
| **Anti-S IgG** | 82.4% | 83.6% | 84.0% | 81.4% |
| **Anti-S IgA or IgG** | 84.9% | 84.3% | 84.7% | 84.7% |

| Assay positivity | weighted by age strata | | | |
|---|---|---|---|---|
| | W2 (N=404) | M1 (N=421) | M3 (N=418) | M6 (N=334) |
| **Anti-S IgA** | 81.3% | 78.1% | 66.6% | 69.6% |
| **Anti-S IgG** | 80.9% | 82.4% | 82.9% | 80.6% |
| **Anti-S IgA or IgG** | 83.4% | 83.1% | 83.6% | 83.8% |

**Table S5**. **Association between demographic and clinical factors and antibody responses over time.** The table demonstrates results from repeated-measures mixed linear regression models assessing the association of demographic and clinical variables with natural logarithm-transformed anti-S-IgA and -IgG mean fluorescence intensity (MFI) ratios in the overall study population (n=431). Models were adjusted for age group, sex, symptom severity based on symptom count and time from diagnosis to testing, using a random intercept for each individual in the study. BMI: Body Mass Index, CI: Confidence Interval, ref: Reference.

| | **log( Anti-S-IgA MFI ratio )** | | **log( Anti-S-IgG MFI ratio )** | |
|---|---|---|---|---|
| **Variable** | **Coefficient (95% CI)** | **P-value** | **Coefficient (95% CI)** | **P-value** |
| **Age group** | | | | |
| 18-39 years | ref | | ref | |
| 40-64 years | 0.34 (-0.02 to 0.69) | 0.0628 | 0.10 (-0.21 to 0.42) | 0.5252 |
| 65+ years | 0.82 (0.46 to 1.17) | <0.001 | 0.78 (0.47 to 1.10) | <0.001 |
| **Sex** | | | | |
| Female | ref | | ref | |
| Male | 0.52 (0.22 to 0.81) | <0.001 | 0.34 (0.08 to 0.60) | 0.011 |
| **Symptom severity (based on symptom count)** | | | | |
| Asymptomatic | ref | | ref | |
| 1-5 symptoms | 0.46 (0.04 to 0.87) | 0.0314 | 0.66 (0.29 to 1.03) | <0.001 |
| ≥6 symptoms | 1.05 (0.64 to 1.46) | <0.001 | 1.28 (0.92 to 1.65) | <0.001 |
| **Hospitalization within first 2 weeks** | | | | |
| Non-hospitalized | ref | | ref | |
| Hospitalized | 0.87 (0.10 to 1.63) | 0.0261 | 0.95 (0.27 to 1.62) | 0.006 |

| | | | | |
|---|---|---|---|---|
| **Smoking status** | | | | |
| Non-Smoker | ref | | ref | |
| Ex-Smoker | -0.06 (-0.41 to 0.30) | 0.7528 | -0.04 (-0.35 to 0.27) | 0.7881 |
| Smoker | -0.43 (-0.85 to 0.00) | 0.0507 | -0.50 (-0.88 to -0.12) | 0.01 |
| **BMI** | | | | |
| Per unit increase | 0.01 (-0.02 to 0.05) | 0.4933 | 0.02 (-0.02 to 0.05) | 0.3718 |
| **Comorbidities** | | | | |
| None | ref | | ref | |
| At least one | -0.02 (-0.36 to 0.32) | 0.9105 | -0.02 (-0.32 to 0.28) | 0.9155 |
| **Immune suppression** | | | | |
| None | ref | | ref | |
| At least one | -0.15 (-1.05 to 0.75) | 0.7478 | -0.28 (-1.07 to 0.52) | 0.4967 |

**Table S6**. **Association between demographic and clinical factors and T cell responses over time.** The table demonstrates results from repeated-measures mixed linear regression models assessing the association of demographic and clinical variables with natural logarithm-transformed total epitope pool-specific T cells in the subsample selected for detailed immune analyses (n=64). Models were adjusted for age group, sex, symptom severity based on symptom count and time from diagnosis to testing, using a random intercept for each individual in the study. BMI: Body Mass Index, CI: Confidence Interval, ref: Reference.

| | **log ( Overall T Cell Count )** | |
|---|---|---|
| **Variable** | **Coefficient (95% CI)** | **P-value** |
| **Age group** | | |
| 18-39 years | ref | |
| 40-64 years | 0.23 (-0.57 to 1.03) | 0.5723 |
| 65+ years | 1.58 (0.70 to 2.46) | <0.001 |
| **Sex** | | |
| Female | ref | |
| Male | 0.11 (-0.57 to 0.79) | 0.7492 |
| **Symptom severity (based on symptom count)** | | |
| Asymptomatic | ref | |
| 1-5 symptoms | 0.73 (-0.19 to 1.64) | 0.1174 |
| ≥6 symptoms | 1.76 (0.94 to 2.58) | <0.001 |
| **Hospitalization within first 2 weeks** | | |
| Non-hospitalized | ref | |
| Hospitalized | -0.08 (-1.18 to 1.03) | 0.8894 |

| | | |
|---|---|---|
| **Smoking status** | | |
| Non-Smoker | ref | |
| Ex-Smoker | 0.98 (0.20 to 1.77) | 0.0151 |
| Smoker | 0.43 (-0.43 to 1.29) | 0.3206 |
| **BMI** | | |
| Per unit increase | -0.01 (-0.09 to 0.07) | 0.88 |
| **Comorbidities** | | |
| None | ref | |
| At least one | 0.42 (-0.41 to 1.25) | 0.3186 |
| **Immune suppression** | | |
| None | ref | |
| At least one | -0.70 (-3.40 to 2.01) | 0.6069 |

**Table S7. Association between demographic and clinical factors and anti-S-IgG antibody positivity at two weeks and six months.** The table demonstrates results from mixed logistic regression models assessing the association of demographic and clinical variables with anti-S-IgG antibody positivity in the overall study population (n=431) at two weeks and six months after diagnosis of SARS-CoV-2 infection. Models were adjusted for age group, sex, symptom severity based on symptom count and time from diagnosis to testing, using a random intercept for each individual in the study. Odds ratios for hospitalization could not be meaningfully estimated since all hospitalized participants were tested antibody positive. BMI: Body Mass Index, CI: Confidence Interval, OR: Odds Ratio, ref: Reference.

| | Anti-S-IgG Positivity at 2 Weeks | | Anti-S-IgG Positivity at 6 Months | |
|---|---|---|---|---|
| **Variable** | **OR (95% CI)** | **P-value** | **OR (95% CI)** | **P-value** |
| **Age group** | | | | |
| 18-39 years | ref | | ref | |
| 40-64 years | 0.85 (0.45 to 1.61) | 0.6245 | 0.89 (0.47 to 1.69) | 0.7242 |
| 65+ years | 1.87 (0.91 to 3.95) | 0.0931 | 1.56 (0.70 to 3.68) | 0.2873 |
| **Sex** | | | | |
| Female | ref | | ref | |
| Male | 1.98 (1.15 to 3.48) | 0.0154 | 1.62 (0.91 to 2.95) | 0.1062 |
| **Symptom severity (based on symptom count)** | | | | |
| Asymptomatic | ref | | ref | |
| 1-5 symptoms | 1.87 (0.97 to 3.62) | 0.0615 | 1.35 (0.63 to 2.83) | 0.4278 |
| ≥6 symptoms | 6.25 (2.94 to 13.72) | <0.001 | 4.23 (1.88 to 9.67) | <0.001 |

| | | | | |
|---|---|---|---|---|
| **Hospitalization within first 2 weeks** | | | | |
| Non-hospitalized | ref | | ref | |
| Hospitalized | n.e. | n.e. | n.e. | n.e. |
| **Smoking status** | | | | |
| Non-Smoker | ref | | ref | |
| Ex-Smoker | 0.62 (0.31 to 1.26) | 0.181 | 0.76 (0.37 to 1.65) | 0.4771 |
| Smoker | 0.24 (0.12 to 0.51) | <0.001 | 0.34 (0.16 to 0.74) | 0.0059 |
| **BMI** | | | | |
| Per unit increase | 0.99 (0.92 to 1.07) | 0.7217 | 0.97 (0.90 to 1.06) | 0.5105 |
| **Comorbidities** | | | | |
| None | ref | | ref | |
| At least one | 0.82 (0.43 to 1.59) | 0.5558 | 1.15 (0.57 to 2.45) | 0.7041 |
| **Immune suppression** | | | | |
| None | ref | | ref | |
| At least one | 0.66 (0.15 to 4.72) | 0.6268 | 0.57 (0.12 to 4.17) | 0.5156 |

**Table S8**. **Association between demographic and clinical factors and overall T cell positivity at two weeks and six months.** The table demonstrates results from mixed logistic regression models assessing the association of demographic and clinical variables with overall epitope pool-specific T cell positivity in the subsample selected for detailed immune analyses (n=64) at two weeks and six months after diagnosis of SARS-CoV-2 infection. Models were adjusted for age group, sex, symptom severity based on symptom count and time from diagnosis to testing, using a random intercept for each individual in the study. Odds ratios for immune suppression could not be meaningfully estimated. BMI: Body Mass Index, CI: Confidence Interval, OR: Odds Ratio, ref: Reference.

| | **Overall T cell Positivity at 2 Weeks** | | **Overall T cell Positivity at 6 Months** | |
|---|---|---|---|---|
| **Variable** | **OR (95% CI)** | **P-value** | **OR (95% CI)** | **P-value** |
| **Age group** | | | | |
| 18-39 years | ref | | ref | |
| 40-64 years | 0.53 (0.12 to 2.11) | 0.3732 | 0.89 (0.24 to 3.27) | 0.8599 |
| 65+ years | 0.96 (0.18 to 5.16) | 0.9616 | 0.74 (0.14 to 4.09) | 0.7193 |
| **Sex** | | | | |
| Female | ref | | ref | |
| Male | 2.17 (0.69 to 7.81) | 0.2023 | 0.89 (0.27 to 2.99) | 0.8414 |
| **Symptom severity (based on symptom count)** | | | | |
| Asymptomatic | ref | | ref | |
| 1-5 symptoms | 2.69 (0.62 to 13.25) | 0.1978 | 0.98 (0.21 to 4.64) | 0.9815 |
| ≥6 symptoms | 2.06 (0.53 to 8.32) | 0.2959 | 4.23 (0.92 to 21.32) | 0.0673 |
| **Hospitalization within first 2 weeks** | | | | |

| | | | | |
|---|---|---|---|---|
| Non-hospitalized | ref | | ref | |
| Hospitalized | n.e. | n.e. | 5.55 (0.44 to 183.45) | 0.2472 |
| **Smoking status** | | | | |
| Non-Smoker | ref | | ref | |
| Ex-Smoker | 1.46 (0.34 to 6.92) | 0.6156 | 6.24 (1.26 to 45.52) | 0.0399 |
| Smoker | 0.37 (0.09 to 1.51) | 0.1643 | 3.01 (0.52 to 24.97) | 0.2455 |
| **BMI** | | | | |
| Per unit increase | 0.95 (0.82 to 1.10) | 0.4685 | 0.92 (0.79 to 1.06) | 0.2402 |
| **Comorbidities** | | | | |
| None | ref | | ref | |
| At least one | 0.90 (0.27 to 2.91) | 0.8558 | 1.27 (0.35 to 4.89) | 0.7164 |
| **Immune suppression** | | | | |
| None | ref | | ref | |
| At least one | n.e. | n.e. | n.e. | n.e. |

**Table S9**. **Reference table for converting anti-S-IgG MFI ratios to BAU/ml (based on Roche Elecsys Anti-SARS-CoV-2 S immunoassay).** The table demonstrates equivalence values between the Luminex-based MFI ratios and BAU/ml based on a commercial antibody test, derived from cross-validation assays.

| **CHUV Anti-S-IgG (MFI Ratio)** | **Roche Elecsys Anti-S-IgG (BAU/ml)** |
|---|---|
| 1 | 1 |
| 4 | 6 |
| **6** | **12** |
| 10 | 30 |
| 25 | 170 |
| 50 | 656 |
| 100 | 2585 |
| 150 | 5795 |
| 200 | 10289 |
